# Association between oral anticoagulant adherence and serious clinical outcomes in patients with atrial fibrillation: A long-term retrospective cohort study

**DOI:** 10.1101/2023.07.18.23292847

**Authors:** Abdollah Safari, Hamed Helisaz, Shahrzad Salmasi, Adenike Adelakun, Mary A. De Vera, Jason G. Andrade, Marc W. Deyell, Peter Loewen

## Abstract

**Background:** Patients with atrial fibrillation (AF) are frequently nonadherent to oral anticoagulants (OACs) prescribed for stroke and systemic embolism (SSE) prevention. We quantified the relationship between OAC adherence and AF clinical outcomes using methods not previously applied to this problem.

**Methods:** Retrospective observational cohort study of incident cases of AF from population-based administrative data over 23 years. The exposure of interest was proportion of days covered (PDC) during 90 days before an event or end of follow-up. Cox proportional hazard models were used to evaluate time to first SSE and the composite of SSE, TIA, or death, and several secondary outcomes.

**Results:** 44,172 patients were included with median follow-up of 6.7 years. For DOACs, each 10% decrease in adherence was associated with a 14% increased hazard of SSE and 5% increased hazard of SSE, TIA, or death. For VKA the corresponding increase in SSE hazard was 3%. Receiving DOAC or VKA was associated with primary outcome hazard reduction across most the PDC spectrum. Differences between VKA and DOAC were statistically significant for all efficacy outcomes and at most adherence levels.

**Conclusions:** Even small reductions in OAC adherence in patients with AF were associated with significant increases in risk of stroke, with greater magnitudes for DOAC than VKA. DOAC recipients may be more vulnerable than VKA recipients to increased risk of stroke and death even with small reductions in adherence. The worsening efficacy outcomes associated with decreasing adherence occurred without the benefit of major bleeding reduction.

## INTRODUCTION

Atrial fibrillation (AF) is the most common chronic arrhythmia, affecting 3% of the world’s population.^1, 2^ Stroke prevention is a cornerstone of AF management because patients with AF are at 5 times higher risk of experiencing a stroke than those without AF, and strokes secondary to AF are more debilitating and more lethal than those due to cerebrovascular disease.^3, 4^ Oral anticoagulants [OACs; vitamin K antagonists (VKAs, typically warfarin), Direct Oral Anticoagulants (DOACs; apixaban, dabigatran, edoxaban and rivaroxaban)] are highly effective for stroke prevention.^5–8^ Unfortunately, it is estimated that 33-50% of patients are nonadherent to their OACs and miss, on average, 30% of their OAC doses.^9, 10^

It is important to quantify the relationship between OAC adherence and clinical outcomes in patients with AF because the clinical consequences of nonadherence are potentially devastating and costly. The few existing studies have treated adherence as a binary exposure (e.g. ≥ 80 or < 80%) and evaluated the effect of adherence over short follow-up periods (< 2 years),^11–16^ despite OAC therapy being lifelong and adherence a continuous spectrum known to vary over time.^17^ Existing studies have typically excluded VKA users because of difficulties in measuring adherence due to its variable dosing regimens. It is conceivable that patients with different levels of adherence who would otherwise be simply categorized as nonadherent experience different clinical event rates, and that nonadherence to VKA vs. DOACs has significantly different clinical consequences due to their different pharmacodynamics.^18–22^ Hence, significant uncertainty remains about the relationship between OAC adherence and the risk of stroke or systemic embolism (SSE) and death.

Given our limited understanding of the multidimensional nature of medication adherence, reliable interventions to improve adherence in patients with AF remain elusive.^9, 23^ A more comprehensive understanding of these phenomena could help identify patients with adherence problems who are most at risk of adverse clinical outcomes and could inform the design of adherence interventions to maximize their impact on stroke and death.

To address these knowledge gaps, our objective was to quantify the relationship between adherence to OACs and SSE, the composite of SSE, transient ischemic attack (TIA), or death, all-cause death, cardiovascular death, SSE or TIA, ischemic stroke, hemorrhagic stroke, major bleeding, and nontraumatic intracranial hemorrhage, and to compare the consequences of nonadherence to VKA to DOACs in terms of these outcomes.

## METHODS

### Study design and setting

This was a retrospective observational cohort study using de-identified patient records from administrative data for the entire population of the province of British Columbia (BC), Canada (∼5 million residents). This included the following databases: Medical Services Plan (MSP) billing for outpatient visits, Discharge Abstract Database (DAD) containing extensive data from hospitalizations, the Consolidation File containing demographics such as sex, place of residence, and registration with the provincial healthcare plan, and the Vital Statistics Database containing date and primary cause of death. Data for all prescription medications dispensed outside of hospital between January 1996 and December 2019 was retrieved from the PharmaNet database and linked to the other datasets. Person-level linkage used a study-specific unique identifier created by the data provider and linkage was performed before the data was released to the investigators. These data sources are used extensively for patient outcomes research.^24^

The study was approved by the University of British Columbia Clinical Research Ethics Board (H17-02420). Study reporting follows the Strengthening the Reporting of Observational Studies in Epidemiology (STROBE) and Reporting of Studies Conducted Using Observational Routinely Collected Health Data Statement for Pharmacoepidemiology (RECORD-PE) extension guides.^25–27^

### Participants

We created an incident cohort of adult patients ≥ 18 years old with non-valvular AF. Using the algorithm validated by Navar et al. with a positive predictive value of 95.7%,^28^ with minor modification for our datasets as previously published^29, 30^ we included individuals who had ≥3 recorded visits in MSP or DAD related to AF or atrial flutter, with at least one being AF-specific (ICD-9 = 427.31; ICD-10 = I48; Supplemental materials). To increase specificity, at least two of the visits had to occur within one year.^28^ The date of the first qualifying AF diagnosis code was specified as the “diagnosis date”.

Next, using Anatomical Therapeutic Chemical (ATC) codes from PharmaNet records, we identified all patients with at least one filled prescription for an OAC (warfarin, dabigatran, apixaban, rivaroxaban, edoxaban) after their AF diagnosis date or within 60 days prior to that date. All other patients were excluded.

Next, patients were excluded for additional reasons: >60 day gap (on average) in plan enrolment during 3 years prior to first OAC prescription after diagnosis date (to preserve sensitivity of subsequent look-back procedures), OAC prescription during 3 years prior to first post-diagnosis date OAC prescription (to establish incident OAC use), indications for OAC other than non-valvular AF (e.g., venous thromboembolism, rheumatic valve disease; Supplemental materials A1) during 3 years prior to first OAC prescription after diagnosis date, < 2 OAC prescription fills (PDC calculation requires 2 or more prescriptions), and available follow-up <3 months after second OAC prescription fill.

Cohort entry (“index date”) occurred on the date of the second OAC prescription fill. Because index date and cohort entry were the same and outcomes and exposures were time-dependent, risk of bias due to immortal time was minimized.^31^ Potential bias from patients having events between diagnosis date (which was before cohort entry) and index date were minimized by including such events as a covariate in study models.

Baseline patient characteristics were measured during 12 months prior to the index date (“baseline period”). Follow-up ended at the time of first study event, gap in plan enrolment of >30 days, or on the date of the last available data, whichever came first.

### Variables

The primary exposure of interest was OAC adherence measured as proportion of days covered (PDC).^32–34^ Ninety-day consecutive windows (the typical length of dispensed prescriptions in the study database) were created for each patient to approximate the average duration of OAC therapy between their first OAC prescription fill date and the end of their follow-up and PDC was calculated for each window.^35^ The validated Random Effects Warfarin Days’ Supply (REWarDS) method was used to estimate the average daily dose for warfarin in the computation of PDC for warfarin.^36^ PDC is a value between zero (total non-adherence) and one (complete adherence). However, to allow more meaningful interpretation of its association with outcomes in our study models, PDC was multiplied by 10, representing PDC deciles, where each unit increase means 10% absolute increase in PDC.

The co-primary outcomes were SSE and the composite of SSE, transient ischemic attack (TIA), or death. Secondary outcomes were all-cause death, cardiovascular death, SSE or TIA, ischemic stroke, hemorrhagic stroke, major bleeding, and nontraumatic intracranial hemorrhage. All outcomes were measured as time to first event.

All outcomes except death were based on the first or second most responsible causes of hospitalization. Death and its cause were ascertained from the Vital Statistics registry. The primary outcome of SSE was chosen to align the Canadian Cardiovascular Society’s AF surveillance quality indicators program’s primary outcome and used the same validated ascertainment scheme.^37, 38^ The other primary outcome was chosen to avoid competing risk between SSE and death and included TIA using a validated ascertainment scheme to encompass all stroke-related outcomes.^39^

Covariates included in study analyses were age, sex, SSE risk score, CHA_2_DS_2_-VASc (Congestive heart failure, Hypertension, Age ≥75, Diabetes, prior Stroke or systemic embolism, Vascular disease, Age 65-74, Sex [female]),^40^ major bleeding risk score, modified HAS-BLED (Hypertension, Abnormal renal/liver function, Stroke, Bleeding, Labile INR, Elderly, and Drugs/alcohol),^41, 42^ comorbidities (individual components of CHA_2_DS_2_-VASc and HAS-BLED scores), socioeconomic status, neighbourhood income quintile, number of medical services plan billings, number of hospitalizations, time from AF diagnosis to OAC start, Charlson and Elixhauser comorbidity indices (CCI, ECI),^43, 44^ and polypharmacy (5 or more concurrent medications, not including OAC).^45^ Following convention, CHA_2_DS_2_-VASc and HAS-BLED scores were binarized at ≥2. Due to their highly skewed distributions, number of hospitalizations during follow-up was grouped into four categories: 0, 1, 2, and ≥3 admissions, and time to OAC initiation was grouped into three categories: <3 months, 3 to 9 months, and >9 months. Multivariable imputation was used to impute missing values for socioeconomic status (SES), the only covariate with missing values.

To enable analyses of association between specific OAC drugs and the study outcomes, we created an additional variable for OAC prescribed at time of event using a novel scheme (Supplemental materials A4). For our main exposure of interest, PDC, to study the effect of OAC adherence and OAC drug class separately, we included their corresponding information in three separate variables: PDC, OAC drug class, and individual OAC drug. The former measured patients’ adherence to any prescribed OAC medication calculated during a 90-day window prior to the event date or end of follow-up, and the others captured the class or drug temporally associated with the event.

Since HAS-BLED, CHA_2_DS_2_-VASc, CCI, and ECI were highly correlated, we included CHA_2_DS_2_-VASc in all study models except for major bleeding, where HAS-BLED was substituted, and CCI and ECI were excluded from all models.

### Statistical analyses

To evaluate the association between OAC adherence and time to clinical outcomes, multivariable Cox proportional hazard models were constructed that incorporated PDC and the study covariates. To ensure strokes and major bleeds (and their subtypes) were incident events related to the patient’s AF, for each model we excluded patients with a history of that event prior to their AF diagnosis date. For example, patients with an ischemic stroke prior to AF diagnosis date were excluded from models with stroke as an outcome. Patients with events occurring between diagnosis date and index date were not excluded and the event was captured in a covariate as explained above. For each model, history of other study outcomes within the previous 3 years was included as a covariate. For example, when stroke was among the model outcomes, history of major bleeding was used as a covariate in the model, and vice-versa. For outcomes involving death, history of stroke and major bleeding were both included in the model. For all analyses we used each patient’s OAC PDC during the 90-days prior to the event or end of follow-up, whichever came first. Sex was included as a covariate in all models regardless of its contribution.

To analyse how associations between PDC and risk of outcome varied based on OAC class (VKA or DOAC) and the specific OAC drug the patient was receiving at the time of their event, we fitted the Cox models with an additional PDC:OAC class interaction term. We also did contrast analyses comparing the PDC hazard ratios (HRs) between VKA and DOAC recipients. Cox models with adherence to individual OAC drugs as covariates were constructed for the primary outcomes using similar methods.

Model results are presented with HRs and 95% confidence intervals (95%CI) for covariates. HRs of PDCs are changes in risk of outcome per 10% absolute increase in PDC. For ease of interpretation, associations between PDC and study outcomes are also presented as percent increase in hazard of outcome per 10% absolute decrease in PDC.

### Sensitivity analyses

To identify period effects, we fitted the models on patients diagnosed after 2010 when both OAC drug classes were available. To evaluate the scheme used to assign patients an OAC drug class at event time, similar Cox models were fitted on only patients with a stable OAC (no recent drug switches or stopped medications), and different gap times (5, 7, or 10 days) prior to event date were used in the scheme to test their effects on classification and outcomes. To study the effect of PDC on outcomes among only patients with an active OAC, we used similar Cox models but excluded patients with no OAC at event date. Further sensitivity analyses were conducted to test the robustness of the results against different pre-event PDC periods: 30 days and 180 days vs. conventional 90-day windows. We evaluated the effects of excluding patients with a study outcome event prior to AF diagnosis (the primary analysis) by running the models on the full cohort without such exclusions. To further explore effects of sex, we constructed separate Cox models for females and males for the primary outcomes. Finally, to identify residual confounding due to healthy adherer effects,^46^ we performed analyses using dental problems and accidental death as falsification endpoints.^47^

All analysis was conducted using R v4.0.5 (R Core Team, Vienna, Austria, 2021) and RStudio v1.3.1039 (Rstudio Team, Boston, US, 2020).

## RESULTS

### Participants

The cohort included 44,172 people; 44.0% were female, and average age was 70.6 (SD 11.4) years. A participant selection flow diagram is in Supplemental materials A0. Median follow-up time was 6.8 years [interquartile range (IQR): 4.1, 10.9]. The median number of OAC prescriptions filled during follow-up was 14 (IQR 7, 26) per patient. Other characteristics of the included patients are shown in Table 1.

During follow up 17,052 SSE, TIA, or death events and 3257 SSE events occurred. Table 2 shows the number of events included and median time to event in each study model.

**Figure 1:**
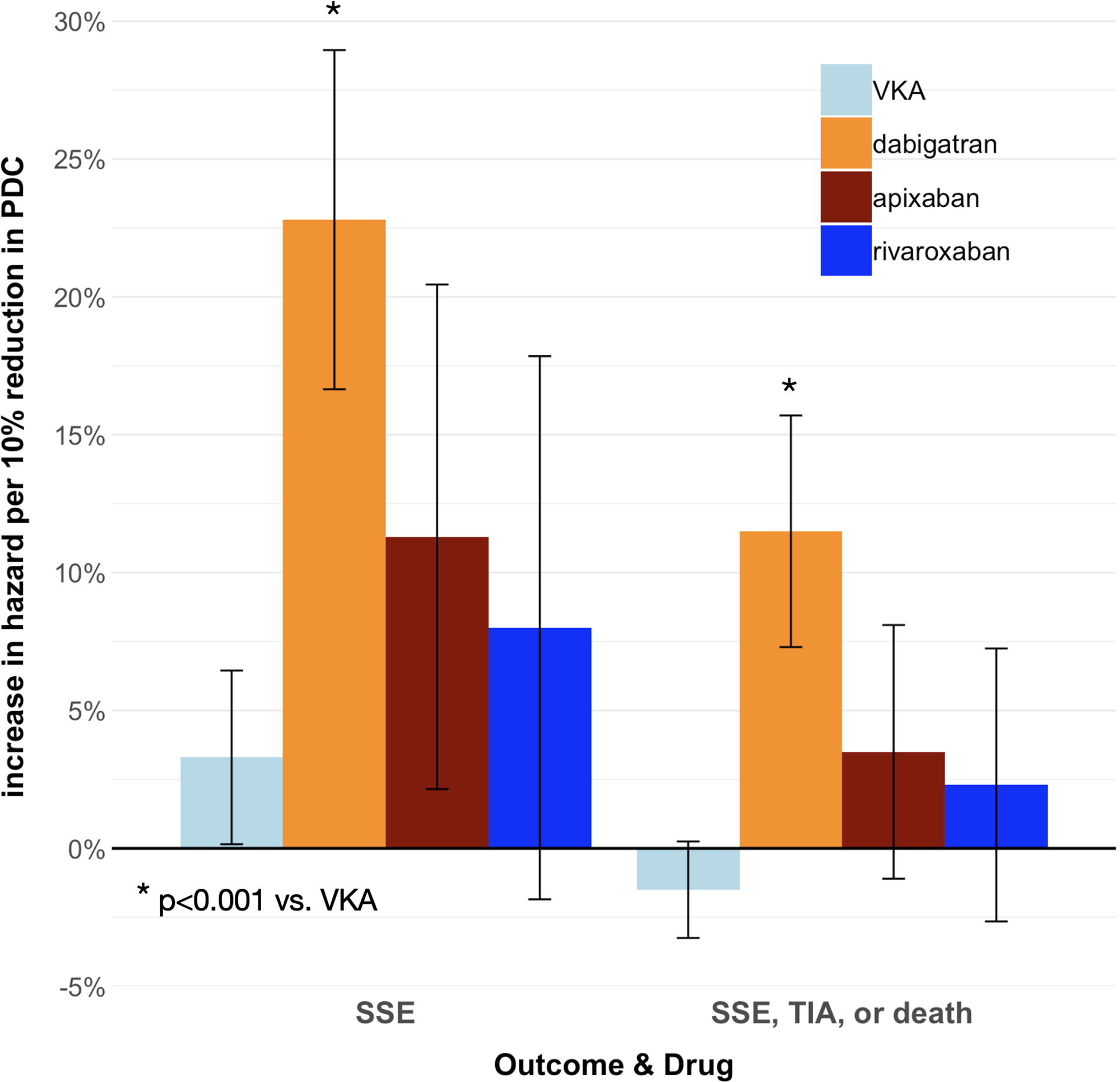
Study design.

### Associations between OAC adherence and clinical outcomes

Results of the time-to-event analyses of the effect of PDC on outcomes are shown in Table 3 and Figure 2. Significant associations between PDC and outcomes were identified for SSE (for VKA and DOACs), SEE, TIA, or death (for DOAC only), SSE or TIA (for DOAC only), and ischemic stroke (for VKA and DOAC). For these outcomes, reductions in PDC were associated with increases in hazard of outcome with significantly larger effects for DOAC than VKA.

**Figure 2:**
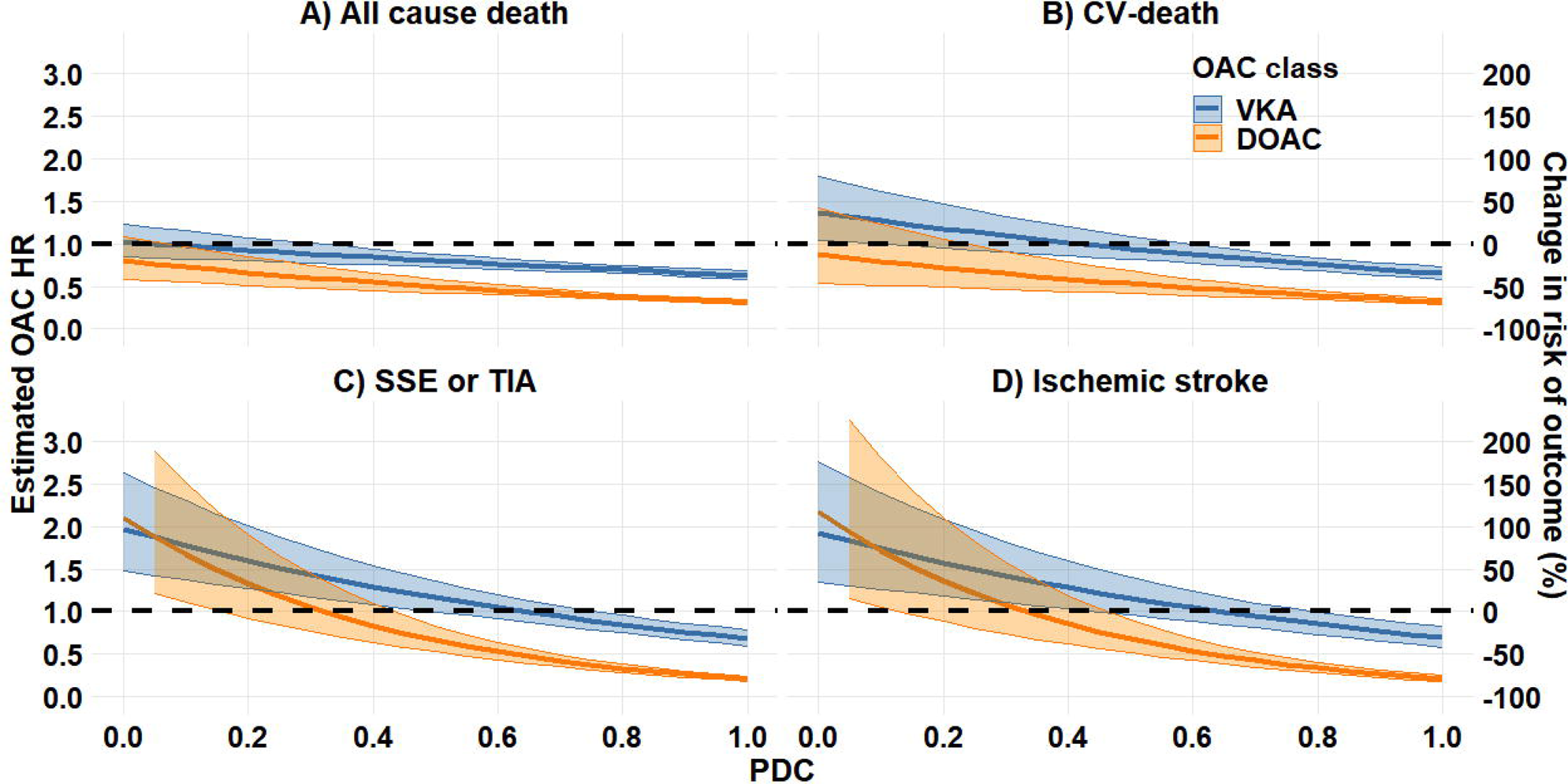
Increase in hazard of clinical event per 10% reduction in PDC by OAC drug class. *VKA vs. DOAC test for contrast p<0.001 SSE: stroke or systemic embolism; TIA: transient ischemic attack; VKA: vitamin K antagonist; DOAC: direct-acting oral anticoagulant; PDC: proportion of days covered

Conversely, lower PDC was associated with reduced hazard of all-cause death for VKA only, and for major bleeding lower PDC for both VKA and DOAC were associated with increased hazard of the outcome.

Figures 3 and 4 depict the change in hazard of event in OAC recipients compared to having no OAC available at time of event for the primary and secondary efficacy outcomes, across the spectrum of PDC. Several observations are warranted: The effect of OAC exposure depended on PDC level, ranging from HR 1.96 to 0.20 (equivalently, from 96% to −80% change in event risk) for SSE, and from HR 1.13 to 0.30 (13% to −70% change in event risk) for SSE, TIA, or death. Being under OAC and hazard of all efficacy outcomes were inversely associated at most PDC levels (blue and orange curves under dashed lines), with greater risk reductions at higher values of PDC and among DOAC recipients. More specifically, DOAC recipients had a significantly lower risk of SSE with PDC >0.4, and >0.2 for SSE, TIA, or death. VKA recipients, however, needed to have higher PDC levels (>0.7 and >0.4, respectively) to be negatively associated with primary outcome risks. Additionally, the negative association between being under OAC and risk of outcomes became stronger (larger OAC effects) as PDC increased for both primary outcomes. These increasing trends of OAC effect slowed down, but persisted, at higher adherence levels through to PDC 1.0. These curves becoming parallel at higher PDC levels >∼0.5 indicates similar amounts of risk reduction for VKA and DOAC per increment in PDC.

**Figure 3:**
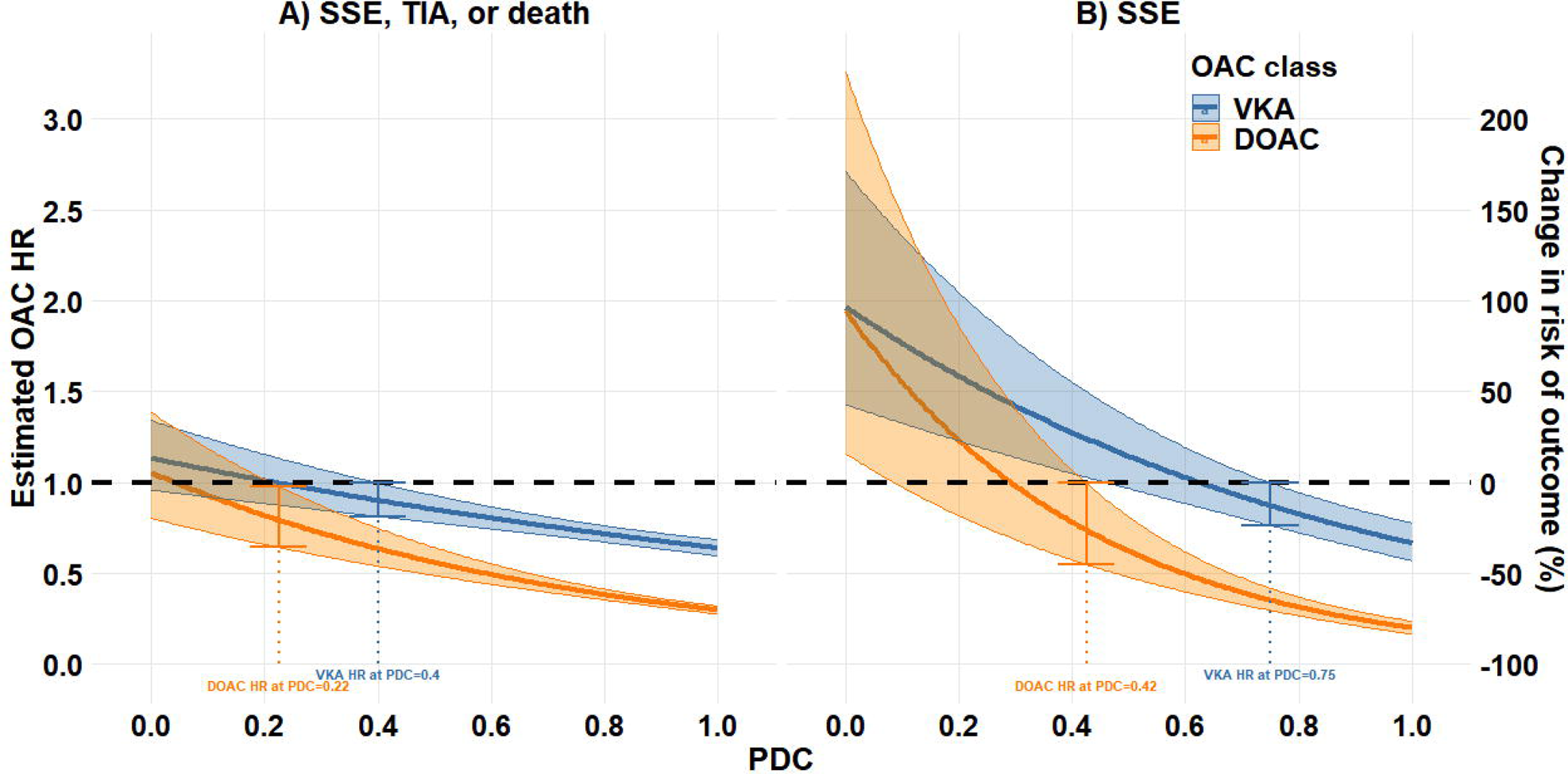
Estimated OAC hazard ratios and change in risk of primary outcomes along with their 95% CI across the PDC spectrum. The black dashed horizontal line corresponds to no effect (HR=1) and the vertical lines indicate the minimum PDC at which being under the corresponding OAC was significantly negatively associated with the risk of outcome. SSE: stroke or systemic embolism; TIA: transient ischemic attack; VKA: vitamin K antagonist; DOAC: direct-acting oral anticoagulant; PDC: proportion of days covered

**Figure 4:**
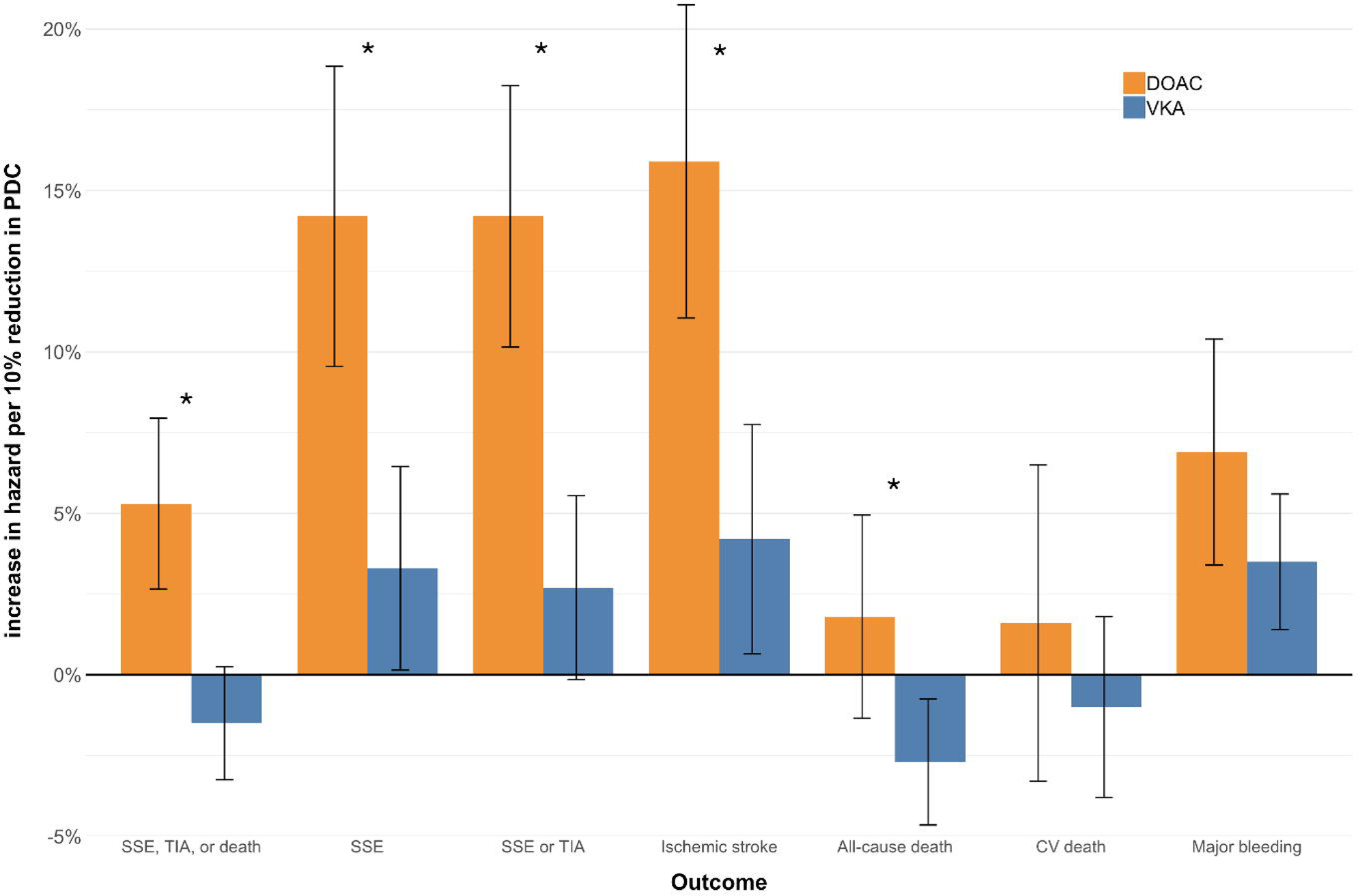
Estimated OAC hazard ratios and change in risk of secondary efficacy outcomes along with their 95% CI across the PDC spectrum. The black dashed horizontal line corresponds to no effect (HR=1). SSE: stroke or systemic embolism; TIA: transient ischemic attack; VKA: vitamin K antagonist; DOAC: direct-acting oral anticoagulant; PDC: proportion of days covered.

DOAC recipients had lower risks of outcomes across the PDC spectrum compared to VKA recipients for both primary outcomes, and a significantly lower risk when PDC levels were greater. The gaps between DOAC and VKA estimated hazards became larger as PDC increased due to faster changes in DOAC effects (larger absolute values of slope), which led to more pronounced differences between VKA and DOAC effects at higher values of PDC.

Conversely, being under OAC with a low adherence (any PDC <0.1 and <0.4 among DOAC and VKA, respectively) was associated with a greater risk of SSE compared to having no OAC available at time of event (estimated HR >1 in leftward portions of Figure 3). Despite apparent lower estimated effect of DOAC than that of VKA at such low PDC levels, the difference between OAC classes was not statistically significant in that PDC range. Neither OAC class was not associated with risk of SSE, TIA, or death at lower values of PDC (dashed line falls within 95% CI of both curves at low PDC in Figure 3).There were too few events to model outcomes for nontraumatic ICH (627 events), and death due to bleeding (372 events), AF (714 events), or stroke (1013 events).

### Individual OAC analyses

Associations between adherence to individual OACs and the primary outcomes are shown in Figure 5. Too few participants were receiving edoxaban to include it in the models. Hazards of reduced PDC were larger for dabigatran, apixaban, and rivaroxaban than for VKA although these were statistically significant only for dabigatran for both primary outcomes. Apixaban vs. VKA had borderline significance (p=0.051) for SSE, TIA, or death.

**Figure 5:**
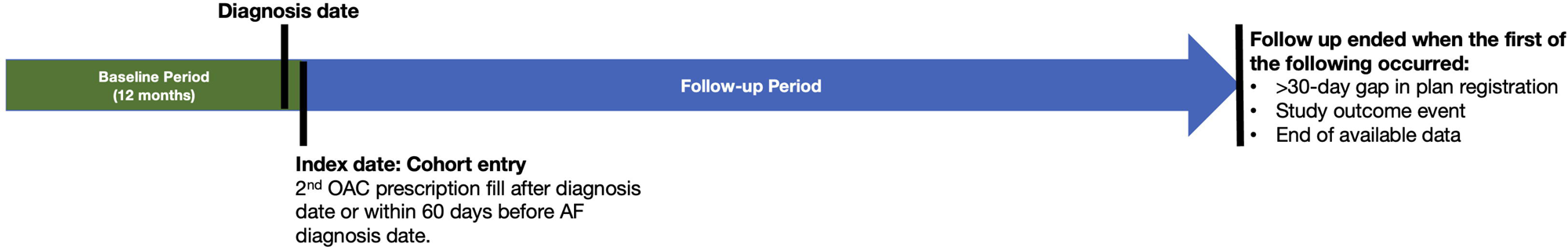
Increase in hazard of clinical event per 10% reduction in PDC by OAC drug. SSE: stroke or systemic embolism; TIA: transient ischemic attack; VKA: vitamin K antagonist; DOAC: direct-acting oral anticoagulant; PDC: proportion of days covered * test for contrast

### Sex-based analyses

Sex-specific Cox models showed similar results for the primary outcomes as in the primary analyses (Supplemental materials Table A6.10). For SSE, TIA, or death, a 10% absolute decrease in PDC for DOAC recipients was associated with a 5.3% and 5.8% increase in hazard for females and males, respectively, and contrast between VKA and DOAC was significant in both sex-specific models (males: p<0.001; females: p=0.014) for SSE, TIA, or death. For SSE, in the females-only Cox model, a 10% absolute decrease in PDC for VKA was associated with a 5.5% increase in hazard of SSE (HR 0.945; 95%CI 0.904 – 0.988), and 17.1% increase in hazard for DOAC recipients (HR 0.829; 95%CI 0.768 – 0.894). The corresponding models for males showed that a 10% absolute decrease in PDC for VKA was not significantly associated with hazard of SSE (HR 0.986; 95%CI 0.939 – 1.036), and 12.2% increase in hazard for DOAC recipients (HR 0.878; 95%CI 0.814 – 0.947). Contrast between VKA and DOAC was significant in both sex-specific models (males: p=0.012; females: p=0.003) for SSE.

### Sensitivity analyses

Confining the primary analyses to patients who initiated OAC therapy after 2010, when at least one alternative to VKA was available, yielded similar model outputs for DOACs and larger effect sizes in the same direction as the main analyses for VKA. Choosing a longer window of PDC prior to events (180 days) resulted in more model instability and slightly reduced effect sizes, indicating clinical events are closely related to short-term OAC-taking behaviours rather than long-term preventive effects, as hypothesized. Sensitivity analysis of the robustness of our scheme to attribute drug classes to clinical events showed that different cut-offs for number of days prior to events for drug changes (5d, 7d, 10d) had minimal effects on the classification proportions. We further tested this scheme by excluding patients who had an OAC change within 7 days fbefore of their event (i.e., stable OAC users only), and found the effect sizes were slightly smaller but with no changes in significance. Exclusion of patients with a stroke event between diagnosis date and index date had negligible effects on the results. The falsification analyses using dental problems and accidental death as outcomes showed no association with PDC for DOAC recipients and small associations in divergent directions for VKA recipients, indicating healthy adherer effects did not contribute significantly to our results. Results of sensitivity analyses are in Supplemental materials A8.

## DISCUSSION

In this large population-based cohort study of incident AF patients with incident OAC use, we found strong associations between OAC adherence and major clinical outcomes. These associations were markedly more pronounced for DOACs than for VKA recipients and applied most strongly to stroke outcomes. For DOACs, each 10% absolute decrease in adherence was associated with a 14.2% increased hazard of SSE and 5.3% increased hazard of SSE, TIA, or death. For VKA the corresponding increase in hazard was significant only for SSE at 3.3%. Similar statistically significant relationships were seen with secondary outcomes of ischemic stroke and SSE or TIA. Across most of the PDC spectrum, being under OAC was associated with SSE and SSE, TIA, or death hazard reduction. The difference in association with these outcomes between DOAC and VKA adherence were most pronounced at PDCs >0.5, and below this level being an OAC recipient yielded similar SSE hazard reduction for both DOAC and VKA.

These results confirm the importance of maximizing adherence in OAC recipients with AF, but especially among DOAC recipients given the much sharper declines in efficacy we observed for nonadherence to DOACs than for VKA. This may be a consequence of the short duration of action of DOACs compared to VKA,^48–50^ with missed DOAC doses potentially leaving patients subtherapeutically anticoagulated for longer periods than missed VKA doses. Our results are the first to demonstrate the magnitude of these differential effects on major clinical outcomes.

Our results also indicate that the conventional binary threshold of 80% for adherence may not be statistically or clinically meaningful, with levels well below that, particularly for DOACs, marking the efficacy boundary in our analyses. We did observe reductions in risk of outcomes, particularly for SSE and other stroke outcomes, at high levels of PDC but 80% did not appear to be an especially meaningful level.

Lower adherence to OACs was associated with higher risk of major bleeding. We conducted some exploratory analyses (e.g., examining only GI bleeding, concurrent NSAID, antiplatelet, or gastroprotective drug therapy, HAS-BLED stratum-specific analyses) to identify potential confounders, but this deserves further study. Other studies have also observed this inverted association between exposure (adherence) and outcome (major bleeding)^51–53^ or lack of association.^54–56^ A plausible explanation is that it is mediated by patient understanding (e.g., via education) about their condition and OAC bleeding risk reduction, factors which also increase OAC adherence and which are variables not available in administrative datasets. Other “healthy adherer” effects are also possible, though our falsification endpoint analyses mitigate this somewhat. The possibility that the relationship between OAC adherence and major bleeding behaves differently than efficacy outcomes deserves focused investigation. Regardless, our results clearly showed that improving OAC adherence does not come at the cost of more major bleeding. Also unexpected was our finding that lower VKA adherence was associated with lower risk of all-cause death, which deserves further investigation.

Importantly, although significant differences were observed between VKA and DOAC adherence’s associations with the study outcomes, this does not prove that the drugs themselves are the cause of the differences. For example, VKA recipients appeared to have had higher stroke and bleeding risk scores as well as more comorbidity than DOAC recipients at baseline. Including these covariates in the models may have reduced bias from OAC prescribing choices but did not fully resolve the issue. There may be other unobserved or unmeasured factors that still confound the treatment effect, and there may be interactions or non-linear effects that are not captured by the covariates. Moreover, including too many covariates may introduce noise or multicollinearity (e.g., between stroke/bleeding scores and comorbidity indices), which can affect the precision and validity of the estimates.^57–60^ Therefore, other methods such as matching, propensity scores, or instrumental variables would be required to address the remaining bias and to infer causality of the differences, a focus of our future work.

Others have demonstrated that poor OAC adherence is associated with worse clinical outcomes in patients with AF.^11–16^ Our work builds upon those findings by applying several methodological techniques rarely used in other OAC adherence studies. We used one of the world’s most comprehensive administrative datasets which includes complete medication prescribing records regardless of payer. We avoided prevalent user biases with a new-user and incident cohort design.^61, 62^ We achieved high statistical power via our large sample size. We used adherence as a continuous variable, revealing the effects of incremental and relatively small (e.g., 10%) changes in adherence, which is not possible when adherence is treated as binary as it is in most studies. Moreover, our methods enabled estimation of the effect of OACs at different levels of PDC and their changes across the PDC spectrum, which also has not been studied before. We used a rigorous scheme to distinguish between OACs in use at the time of the event which allowed us to compare effect estimates between OACs. We included VKA recipients, a group routinely excluded from population-based studies due to difficulties in accurately ascertaining PDC. Finally, our data covered through the end of 2019, which coincided with the beginning of the COVID-19 pandemic and resulted in major changes to prescription filling and drug coverage for OACs in the study jurisdiction. Hence, our results were not confounded by those disruptions.

### Limitations

These results should be interpreted considering the study’s limitations. We assumed that filling a prescription equates to consuming the medication, which may not always be true. As with all observational studies, residual confounding was possible, particularly from some variables known to affect adherence (e.g., education level, adverse effects experienced, psychosocial factors) which were absent from the administrative databases, limiting our ability to account for their contributions. Requiring at least two prescription fills for inclusion in the cohort meant that we excluded those who never initiated therapy or stopped after one prescription fill, a potentially high-risk subpopulation. This concern is mitigated by the fact that patients excluded for filling only one OAC prescription had less comorbidity and stroke risk than our study cohort (Supplemental materials Table A8). The available data did not include laboratory values, so we were unable to validate VKA adherence with INR values or include measures of renal function in study models. Even if INR data were available, we would have required a common adherence variable between DOACs and VKA, and our use of an accurate validated model to ascertain warfarin days supply for individual patients provided reliable PDC measures for VKA and DOAC recipients. There were too few edoxaban recipients and too few of certain events (ICH, deaths due to bleeding, AF, or stroke) to permit analyses. As mentioned, residual confounding may have influenced our major bleeding results and further analyses of the relationship between OAC adherence and major bleeding are warranted, particularly since others have found diverging results with this outcome, as discussed above. Nonetheless, it is doubtful that reduced adherence to OACs within the ranges studied is associated with significantly less major bleeding, the threshold for which may be lower than the thresholds for efficacy outcomes, a phenomenon we are currently studying. A further limitation is that the relationship between adherence and outcomes may not be linear across the PDC spectrum, as our models dictated. This also deserves further study. We were unable to distinguish clinician-guided therapy discontinuations (temporary or permanent) from patient-initiated ones in our calculation of PDC. Considering the life-long nature of thromboprophylaxis in AF patients, the few absolute contraindications to OACs, and consensus guidelines recommending they be resumed even after major bleeding,^63–65^ we believe this to have had a minor impact in our estimation of OAC adherence.

## CONCLUSIONS

Even small reductions in OAC adherence in patients with AF were associated with significant increases in risk of SSE and ischemic stroke, the magnitudes of which were significantly greater for DOAC than VKA. Across most of the PDC spectrum, receiving OAC was associated with SSE hazard reduction. Changes in OAC effects on outcomes were most pronounced at PDCs <0.5 for both DOAC and VKA recipients, and above this level the differences between OAC classes in risk reduction were most pronounced. The conventional adherence threshold of 80% did not appear meaningful in our analyses. These results suggest that DOAC recipients may be more vulnerable than VKA recipients to increased risk of stroke and death even with small reductions in adherence. The worsening efficacy outcomes associated with decreasing adherence occurred without the benefit of major bleeding reduction.

## Acknowledgements

The authors are grateful to Dr. Anita I. Kapanen from University of British Columbia for her expert support of the data stewardship and ethical oversight of this study.

## Data Availability Statement

Access to data provided by the Data Stewards is subject to approval but can be requested for research projects through the Data Stewards or their designated service providers. The following data sets were used in this study: Medical Services Plan (MSP), Discharge Abstract Database (DAD), Consolidation File, Vital Statistics Database, and PharmaNet. You can find further information regarding these data sets by visiting the PopData project webpage at: https://my.popdata.bc.ca/project_listings/17-149/collection_approval_dates. All inferences, opinions, and conclusions drawn in this publication are those of the author(s), and do not reflect the opinions or policies of the Data Steward(s).

