## Supplemental materials for "Association between oral anticoagulant adherence and serious clinical outcomes in patients with atrial fibrillation: A long-term retrospective cohort study"

### SUPPLEMENTAL APPENDICES

#### Participant selection flow diagram

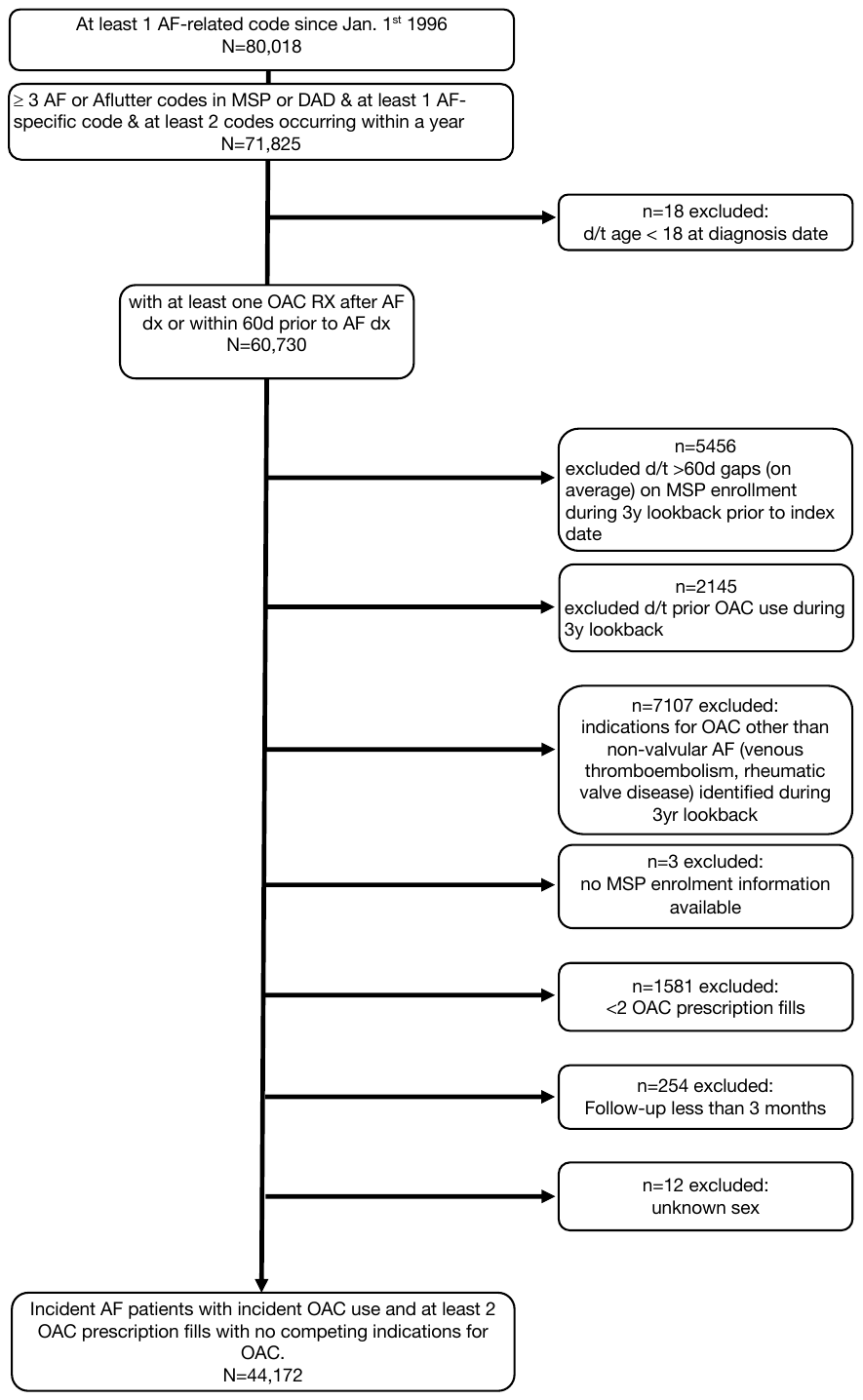

#### Table A1: International Classification of Diseases (ICD) codes used for cohort definitions, outcome definitions, and for calculating risk scores

|  | ICD-9 | ICD-10 |
| --- | --- | --- |
| Qualifying AF ICD codes |  |  |
| Cardiac Dysrhythmia | 427 |  |
| Atrial Fibrillation and Flutter | 427.3 | I48 |
| Atrial Fibrillation | 427.31 | I48.0-2, I48.9 |
| Atrial Flutter | 427.32 | I48.3, I48.4 |
| Excluding AF ICD codes |  |  |
| Pulmonary embolism | 415.1 | I26 |
| Phlebitis and thrombophlebitis | 451 | I80 |
| Other venous embolism and thrombosis | 453 |  |
| Disease of mitral and aortic valve | 394,395,395 | I05, I08 |
| Other rheumatic heart diseases | 398 | I06, I09 |
| Budd-Chiari syndrome |  | I82 |
| Exposure for stroke risk calculation: CHA_2_DS_2_-VASc | | |
| **C**ongestive Heart failure | 398.91, 402.01, 402.11, 402.91, 404.01, 404.03, 404.11, 404.13, 404.91, 404.93, 428.x | I50, I11.0, I13.0, I13.2, I42.0 |
| **H**ypertension | 401.x, 402.x, 403.x, 404.x, 405.x, 437.2 | I10-I13, I15 |
| **A**ge¶ | N/A | N/A |
| **D**iabetes | 250.x, 357.2, 362.0, 366.41 | E10, E11, E13, E14 |
| **S**troke/Transient Ischemic Attack | 433.xx, 434.xx, 435.x, 431, 436, 438.x | I63, I64, G45, I69, I74 |
| **Va**scular disease | 410.x, 411.x, 412.x, 413.x, 414.x, 440.x, 447.1, 557.1, 557.9 | I21, I23, I252, I70–73 |
| **S**ex **c**ategory | N/A | N/A |
| Exposure for bleeding risk calculation: Modified† HAS-BLED | | |
| **H**ypertension‡ | 401, 402, 403, 404, 405 | I10-I13, I15 |
| **A**bnormal renal function  (Kidney disease) | 403.01, 403.11, 403.91, 404.02, 404.03, 404.12, 404.13, 404.92, 404.93, 582, 583.0, 583.1, 583.2, 583.3, 583.4, 583.5, 583.6, 583.7, 585, 586, 588.0, V42.0, V45.1, V56 | I12, I13, N00, N01, N02, N03, N04 N05, N07, N11, N14, N17, N18, N19, Q61 |
| **A**bnormal liver function  (Liver disease) | 070.22, 070.23, 070.32, 070.33, 070.44, 070.54, 070.6, 070.9, 456.0, 456.1, 456.2, 570, 571, 572.2, 572.3, 572.4, 572.5, 572.6, 572.7, 572.8, 573.3, 573.4, 573.8, 573.9, V42.7 |  |
| **S**troke or Transient Ischemic Attack | 433.xx, 434.xx, 435.x, 431, 436, 438.x | I63, I64, G45, I69, I74 |
| **B**leeding history (Major) | 280.0, 285.1, 423.0, 430, 431, 432.x, 455.2, 455.5, 455.8, 459.0, 456.0, 456.20, 459.0, 530.21, 530.7, 530.82, 531.0x, 531.2x, 531.4x, 531.6x, 532.0x, 532.2x, 532.4x, 532.6x, 533.0x, 533.2x, 533.4x, 533.6x, 534.0x, 534.2x, 534.4x, 534.6x, 535.01, 535.11, 535.21, 535.31, 535.41, 535.51, 535.61, 535.71, 537.83, 537.84, 562.02, 562.03, 562.12, 562.13, 568.81, 569.3, 569.85, 578.x  852.x, 853.x  596.7, 599.7, 719.1x, 784.7, 784.8, 786.3 | I60, I61, I62, K250, K252, K254, K260 K262, K264, K270, K272, K274, K280, K282, K290, K920, K921, K922, D62, J942, H113, H356, H431, N02, R04, R31, R58 |
| **L**abile INR (NA – modified HAS-BLED) | N/A | N/A |
| **E**lderly >65 years old ¶ | N/A | N/A |
| **D**rug§ or  Alcohol use  (Alcoholism) | 265.2, 291.1, 291.2, 291.3, 291.5, 291.6, 291.7, 291.8, 291.9, 303.0, 303.9, 305.0, 357.5, 425.5, 535.3, 571.0, 571.1, 571.2, 571.3, 980, V11.3 | E224, E529A, F10, G312, G621, G721 I426, K292, K70, K860, L278A, O354 T51, Z714, Z721 |
| ICD: International Classification of diseases, ATC: Anatomical Therapeutic Chemical  † Modified to omit labile INR, which isn’t ascertainable from Population Data British Columbia.  ¶ Calculated at time of first OAC prescription  ‡ To overcome the under-coding of hypertension in Population Data British Columbia we utilized alternative coding to detect hypertension cases, a method used previously in this particular database. Subjects with hypertension were identified from combination treatment with at least two classes of antihypertensive drugs.  § non-steroidal anti-inflammatory (NSAIDs) and antiplatelet drugs. ATC codes used are in Table A5. | | |

#### Table A2: ATC codes for drug identification in PharmaNet

| Drug | Anatomical Therapeutic Chemical (ATC) code |
| --- | --- |
| OAC | warfarin: B01AA03  apixaban: B01AF02  dabigatran: B01AE07  edoxaban: B01AF03  rivaroxaban: B01AF01 |
| NSAID | M01A |
| antiplatelets | dipyridamole: B01AC07  ticagrelor: B01AC24  clopidogrel: B01AC04  ticlopidine: B01AC05  prasugrel: B01AC22  aspirin: B02BA or B01AC06 |
| gastroprotective | any gastroprotective drug: A02  proton-pump inhibitors (PPIs): A02B |

#### Table A3: Study outcome definitions

| Outcome  Hospitalization for: | Data source | ICD-10-CA code scheme | Justification | Citations |
| --- | --- | --- | --- | --- |
| SSE (stroke or systemic embolism) PRIMARY | DAD | H34.1, H34.2, I63, I64 (Ischemic stroke)  I74 (Systemic embolism)  I60, I61 (Hemorrhagic Stroke)    in DAD DIAG1 or DIAG2 positions | CCS Canadian AF surveillance approach [Canadian Quality Indicators for Atrial Fibrillation and Atrial Flutter]   - Developed and validated based on Canadian DADs. - Uses “primary and secondary diagnoses”, which we translate as “in DIAG1 or DIAG2” positions in DAD - TIA (G45) not included d/t “lack of specificity” | Wilton SB, et al. Surveillance for Outcomes Selected as Atrial Fibrillation Quality Indicators in Canada: 10-Year Trends in Stroke, Major Bleeding, and Heart Failure. CJC Open. 2021;3(5):609–18.  Which is based on:  Sandhu RK, et al. An Update on the Development and Feasibility Assessment of Canadian Quality Indicators for Atrial Fibrillation and Atrial Flutter. Cjc Open. 2019;1(4):198–205. [**https://www.sciencedirect.com/science/article/pii/S2589790X1930037X**](https://www.sciencedirect.com/science/article/pii/S2589790X1930037X) and  Quan H, et al.. Coding Algorithms for Defining Comorbidities in ICD-9-CM and ICD-10 Administrative Data. Med Care. 2005;43(11):1130–9. [**https://journals.lww.com/lww-medicalcare/Abstract/2005/11000/Coding_Algorithms_for_Defining_Comorbidities_in.10.aspx**](https://journals.lww.com/lww-medicalcare/Abstract/2005/11000/Coding_Algorithms_for_Defining_Comorbidities_in.10.aspx) |
| Sub-outcome:  Ischemic stroke | DAD | H34.1, H34.2, I63, I64  in DAD DIAG1 or DIAG2 positions | Same as for SSE    I63 has PPV ≥82% in most studies; I64 has PPV ≥75% in most studies [McCormack et al] | Same as above, plus  McCormick N, Bhole V, Lacaille D, Avina-Zubieta JA. Validity of Diagnostic Codes for Acute Stroke in Administrative Databases: A Systematic Review. Plos One. 2015;10(8):e0135834. [**https://journals.plos.org/plosone/article?id=10.1371/journal.pone.0135834**](https://journals.plos.org/plosone/article?id=10.1371/journal.pone.0135834) |
| Sub-outcome:  Hemorrhagic stroke | DAD | I60, I61  in DAD DIAG1 or DIAG2 positions | Same as for SSE | Same as above |
| SSE or TIA | DAD | G45 (G45.4 excluded) to the SSE definition above  in DAD DIAG1 or DIAG2 positions | Adds TIA (G45, excluding G45.4) to the CCS SSE definition above.  Leong et al. used “G45.X (exc. G45.4), H34.0, H23.1, I63.X (exc. I63.6), I64.X, I67.6”. We added G45 (excl G45.4) to preserve fidelity to the CCS definition.  Adds ~1000 cases to this outcome. | Leong M, et al. Regional Variation in Transient Ischemic Attack and Minor Stroke in Alberta Emergency Departments. Stroke. 2020;51(6):1820–4. [**https://www.ahajournals.org/doi/10.1161/STROKEAHA.119.027960**](https://www.ahajournals.org/doi/10.1161/STROKEAHA.119.027960) |
| SEE, TIA, or Death  CO-PRIMARY | DAD, VS | SEE or TIA definition above + Death definition below | Top-level composite outcome. |  |
| Major bleeding | DAD | H35.6, H43.1, I60, I61, I62, I67.1, I85.x1, K22.11, K22.6, K25.0, K25.2, K25.4, K25.6, K26.0, K26.0, K26.2, K26.4, K26.6, K27.0, K27.2, K27.4, K27.6, K28.0, K28.2, K28.4, K28.6, K29.x1,K31.80, K55.21,K62.5, K66.1, K92.0, K92.1, K92.2, M25.0, N02, R04, R31, R58  in DAD DIAG1 or DIAG2 positions | Same as for SSE | Same as above, except I67.1 added to conform with Sangal’s validated coding scheme for ICH (a sub-outcome below) |
| Sub-outcome:  Nontraumatic intracranial hemorrhage (ICH) | DAD | I60.X - Nontraumatic subarachnoid hemorrhage  I61.X - Nontraumatic intracerebral hemorrhage  I62.X - Other and unspecified nontraumatic intracranial hemorrhage  I67.1 - Cerebral aneurism, nonruptured  in DAD DIAG1 or DIAG2 positions | specificity 0.83 (0.81 - 0.85); sensitivity 0.89 (0.85 - 0.92) | Sangal RB, et al. Identification of Patients with Nontraumatic Intracranial Hemorrhage Using Administrative Claims Data. J Stroke Cerebrovasc Dis. 2020;29(12):105306. [**https://www.ncbi.nlm.nih.gov/pmc/articles/PMC7686163/**](https://www.ncbi.nlm.nih.gov/pmc/articles/PMC7686163/) |
| Death (all-cause) | VS | N/A – directly ascertained |  |  |
| Sub-outcome:  CV Death | VS | N/A – directly ascertained |  |  |
| Sub-outcome:  AF Death | VS | N/A – directly ascertained |  |  |
| Sub-outcome:  Stroke Death | VS | N/A – directly ascertained |  |  |
| Sub-outcome:  Bleeding Death | VS | N/A – directly ascertained |  |  |

#### Table A4: Assignment of OACs to clinical events

To enable analyses of association between specific OAC drugs and the study outcomes, we created an additional variable for “OAC at time of the event” based on the scheme below. It includes thresholds for recency of starting, stopping, or changing OACs before events to assign attributions of OACs (or lack thereof) to individual clinical events. This approach was based on evidence that AF-associated events such as stroke (all types) and major bleeding are most affected by the therapy (or lack thereof) present in close proximity (i.e., a few days) before events, rather than reflective of longer-term residual effects of OACs.^51, 57-59^

**variables**

“OAC identity change” = Rx for different OAC than the one previously filled (i.e. OAC switch), OR new OAC Rx while patient had no OAC days supply (i.e. new OAC started), OR days supply of OAC ended with no new OAC Rx to replace (i.e. OAC stopped). Does not apply to OAC *dose* changes.

[changeClass] = type of OAC identity change within 7d of event

Possible values:

- no OAC at time of event
- VKA start
- DOAC start
- DOAC-DOAC switch
- VKA-DOAC switch
- DOA-VKA switch
- VKA stop
- DOAC stop

[eventDay] = Day 0. Date of hospitalization for event

[changeDay] = number of days before [eventDay]

[OAC1] = class/drug available on [changeDay]-1

[OAC2] = class/drug available after [changeDay]

[eventOACclass] = OAC class assigned to event for analysis purposes

Possible values:

- no OAC at time of event
- VKA
- DOAC

[eventOACdrug] = OAC drug assigned to event for analysis purposes

Possible values:

- no OAC at time of event
- warfarin
- dabigatran
- rivaroxaban
- apixaban
- edoxaban

**scenarios**

*stable OAC pre-event*

If [changeClass] == “no OAC at time of event”

[eventOACclass] ← OAC class available the day before the event AND [eventOACdrug] ← drug available the day before the event

*stopped DOAC*

for all STROKE and DEATH outcomes:

If [changeClass] == “DOAC stop” AND [changeDay] ≤ 4

[eventOAC] ← DOAC/drugname available 1 day before [changeDay]

ELSE

[eventOACclass] ← no OAC at time of event AND [eventOACdrug] ← no OAC at time of event

RATIONALE: DOAC effect is gone 2d after last dose + 2d to develop thrombus. If DOAC stopped after this, assign DOAC to the event. Ref: Thrombosis Canada & CCS AF 2020 guidelines re: peri-procedural thromboprophylaxis.

for all BLEEDING outcomes:

If [changeClass] == “DOAC stop” AND [changeDay] ≤ 2

[eventOAC] ← DOAC/drugname available 1 day before [changeDay]

ELSE

[eventOACclass] ← no OAC at time of event AND [eventOACdrug] ← no OAC at time of event

RATIONALE: For BLEEDS, > around 48h after stopping DOAC a bleed should not be attributed to the drug

*stopped VKA*

for all STROKE and DEATH outcomes:

If [changeClass] == “VKA stop” AND [changeDay] ≤ 7

[eventOACclass] ← VKA AND [eventOACdrug] ← warfarin

ELSE

[eventOACclass] ← no OAC at time of event AND [eventOACdrug] ← no OAC at time of event

RATIONALE: VKA effect is gone ~5d after last dose + 2d to develop thrombus. If VKA stopped after this, assign VKA to the event. Ref: Thrombosis Canada & CCS AF 2020 guidelines re: peri-procedural thromboprophylaxis.

for all BLEEDING outcomes:

If [changeClass] == “VKA stop” AND [changeDay] ≤ 5

[eventOACclass] ← VKA AND [eventOACdrug] ← warfarin

ELSE

[eventOACclass] ← no OAC at time of event AND [eventOACdrug] ← no OAC at time of event

RATIONALE: For BLEEDS, > ~5d after stopping DOAC a bleed should not be attributed to the drug

*switched VKA to DOAC*

If [changeClass] == “VKA to DOAC” AND [changeDay] ≤ 2

[eventOACclass] ← VKA AND [eventOACdrug] ← warfarin

ELSE

[eventOACclass] ← DOAC AND [eventOACdrug] ← drugname

RATIONALE: after 2d of DOAC patient is stabilized regardless of prior OAC. If change was <2d pre-event, assign the pre-change OAC (VKA). Possible for insufficient washout of VKA prior to starting DOAC exists. It is difficult to decide how to attribute the event in that case, but this method defaults to DOAC in that situation.

*switched DOAC to VKA*

If [changeClass] == “DOAC to VKA” AND [changeDay] ≤ 7

[eventOACclass] ← DOAC AND [eventOACdrug] ← drugname

ELSE

[eventOACclass] ← VKA AND [eventOACdrug] ← warfarin

RATIONALE: INR should be >2 in almost all patients by day 7. If change was <7d pre-event, assign the pre-change OAC.

*switched DOAC to DOAC*

If [changeClass] == “DOAC to DOAC” AND [changeDay] ≤ 2

[eventOACclass] ← DOAC AND [eventOACdrug] ← OAC1 drugname

ELSE

[eventOACclass] ← DOAC AND [eventOACdrug] ← OAC2 drugname

RATIONALE: after 2d on new DOAC patient is stabilized. If change was <2d pre-event, assign the pre-change OAC.

*recent DOAC start*

If [changeClass] == “DOAC start ” AND [changeDay] ≤ 2

[eventOACclass] ← no OAC at time of event AND [eventOACdrug] ← no OAC at time of event

ELSE

[eventOACclass] ← DOAC AND [eventOACdrug] ← OAC2drugname

RATIONALE: after 2d on new DOAC, patient is stabilized. If change was <2d pre-event, assign no therapy.

*recent VKA start*

If [changeClass] == “VKA start ” and [changeDay] ≤ 7

[eventOACclass] ← no OAC at time of event AND [eventOACdrug] ← no OAC at time of event

ELSE

[eventOACclass] ← VKA AND [eventOACdrug] ← warfarin

RATIONALE: INR should be >2 in almost all patients by day 7. After 2d on new DOAC patient is stabilized. If change was <2d pre-event, assign no therapy.

If multiple OACs are available to the patient on day of event:

[eventOACclass] ← most recently filled OAC class AND [eventOACdrug] ← most recently filled OAC drug

#### Tables A5: Count, Rate, and Time to Clinical Events

##### Table A5.1: Primary outcomes

| Event | Count | Annual rate | Time to event  Mean (yr) | Time to event  Median (yr) |
| --- | --- | --- | --- | --- |
| Death (all cause) or Stroke (SSE or TIA) | 17052 | 0.053 | 6.18 | 5.29 |
| Stroke (SSE) | 3257 | 0.010 | 5.11 | 4.07 |

##### Table A5.2: Stroke outcomes

| Event | Count | Annual rate | Time to event  Mean (yr) | Time to event  Median (yr) |
| --- | --- | --- | --- | --- |
| SSE or TIA | 4051 | 0.013 | 5.03 | 4.00 |
| Ischemic | 2588 | 0.008 | 5.11 | 4.08 |
| Hemorrhagic | 442 | 0.001 | 5.20 | 4.19 |

##### Table A5.3: Bleeding outcomes

| Event | Count | Annual rate | Time to event  Mean (yr) | Time to event  Median (yr) |
| --- | --- | --- | --- | --- |
| Major bleeding | 6190 | 0.021 | 4.65 | 3.61 |
| Nontraumatic intracranial hemorrhage (ICH) | 627 | 0.002 | 5.04 | 3.94 |

##### Table A5.4: Death outcomes

| Event | Count | Annual rate | Time to event  Mean (yr) | Time to event  Median (yr) |
| --- | --- | --- | --- | --- |
| All cause | 15785 | 0.046 | 6.70 | 5.87 |
| CVS | 6766 | 0.024 | 7.86 | 6.09 |
| AF | 714 | 0.003 | 7.36 | 6.56 |
| Stroke | 1013 | 0.004 | 6.96 | 6.39 |
| bleed | 372 | 0.002 | 6.48 | 5.75 |

#### Table A6: Study models

In models with no interaction term between PDC and OAC drug class (Tables A6.1 and A6.2), it is straightforward to interpret the estimated HR of the PDC (and OAC drug class) main effect. That is, the exponential of the estimated coefficient of PDC can be interpreted as the overall HR estimate of PDC and can be interpreted accordingly. In models with an interaction term between PDC and OAC drug class (Tables A6.3-A6.10), however, since there is no single overall HR estimate for PDC, one needs to derive HR estimate of PDC for each drug class (VKA and DOAC vs. no OAC at time of event). More specifically, there are two terms in these models involving PDC: 1. PDC main effect (called PDC), and 2. interaction between PDC and OAC drug class (called PDC : OAC drug class). Since OAC drug class has three levels of DOAC, VKA, and no OAC at time of event (reference level), two dummy variables corresponding to the OAC drug class appeared in these models: 1. VKA (1 when patient is under VKA, 0 otherwise), and 2. DOAC (1 when patient is under DOAC, 0 otherwise), where these dummy variables enable us to compare patients under each OAC drug class of VKA and DOAC with the patients in no OAC at time of event group. Therefore, three terms appeared in each model outcome involving PDC:

1. PDC: estimate effect of PDC for patients in no OAC at time of event group of OAC drug class
2. PDC : VKA : estimate the difference between the effect of PDC for patients under VKA against patients in the no OAC at time of event group of OAC drug class
3. PDC : DOAC : estimate the difference between the effect of PDC for patients under DOAC against patients in no OAC at time of event group of OAC drug class

The first term with no further calculations estimates the effect of PDC for patients in no OAC at time of event group of OAC drug class. To estimate the PDC effect for patients under VKA and DOAC, we need to follow these steps:

a. Compute log of estimated HR of PDC
b. Compute log of estimated HR of PDC : VKA (or PDC : DOAC)
c. Add up the two computed log-HRs in parts a and b
d. Exponentiate of the computed sum in part c is the estimated PDC HR for patients under VKA (DOAC)
e. To estimate the SE of the estimated PDC HR, one can easily obtain that via the delta method (by using the model estimated coefficients as well as their estimated var-cov matrix)

For instance, for the model presented in Table A6.3, the estimated PDC HR for each level of OAC drug class will be as follows:

- PDC_no OAC at time of event: 1.07 (95% CI: 1.07-1.08)
- PDC_VKA: 1.01 (95% CI: 0.99-1.03)
- PDC_DOAC: 0.95 (95% CI: 0.92-0.97)

One can follow similar steps as above (steps a-e) to obtain estimated OAC drug class HR for patients under DOAC and VKA (against patients in no OAC at time of event group of OAC drug class) as well. The only difference is, given the continuous nature of PDC, we can estimate the OAC drug class HRs (as HR_VKA and HR_DOAC) for a given fixed value of PDC. For instance, for the model presented in Table A6.3, the estimated OAC drug class HR three different values of PDC will be as follows:

i. PDC = 0
   - HR_VKA: 1.27 (95% CI: 1.12-1.44)
   - HR_DOAC: 1.14 (95% CI: 0.89-1.45)
ii. PDC = 50%
    - HR_VKA: 1.11 (95% CI: 1.04-1.18)
    - HR_DOAC: 0.73 (95% CI: 0.65-0.82)
iii. PDC = 100%
     - HR_VKA: 0.96 (95% CI: 0.92-1.02)
     - HR_DOAC: 0.47 (95% CI: 0.44-0.49)

Hence these HR estimates follow our expectation. More specifically, being under DOAC (VKA) is associated with a lower risk of outcome relative to patients in no OAC at time of event group of OAC drug class if the patient adherence level is at least 50% (around 100%). Otherwise, either the difference between the effect DOAC or VKA (compared to the no OAC at time of event group of OAC drug class) is mostly not significant, or occasionally, it can even be associated with a higher risk of outcome. This result may be due to the patients in the no OAC at time of event group of OAC drug class (i.e., under no active OAC medication at event time) being generally healthier that patients who were taking their medications at event time.

##### Table A6.3: Cox proportional hazards model of time to first death, SSE, or TIA

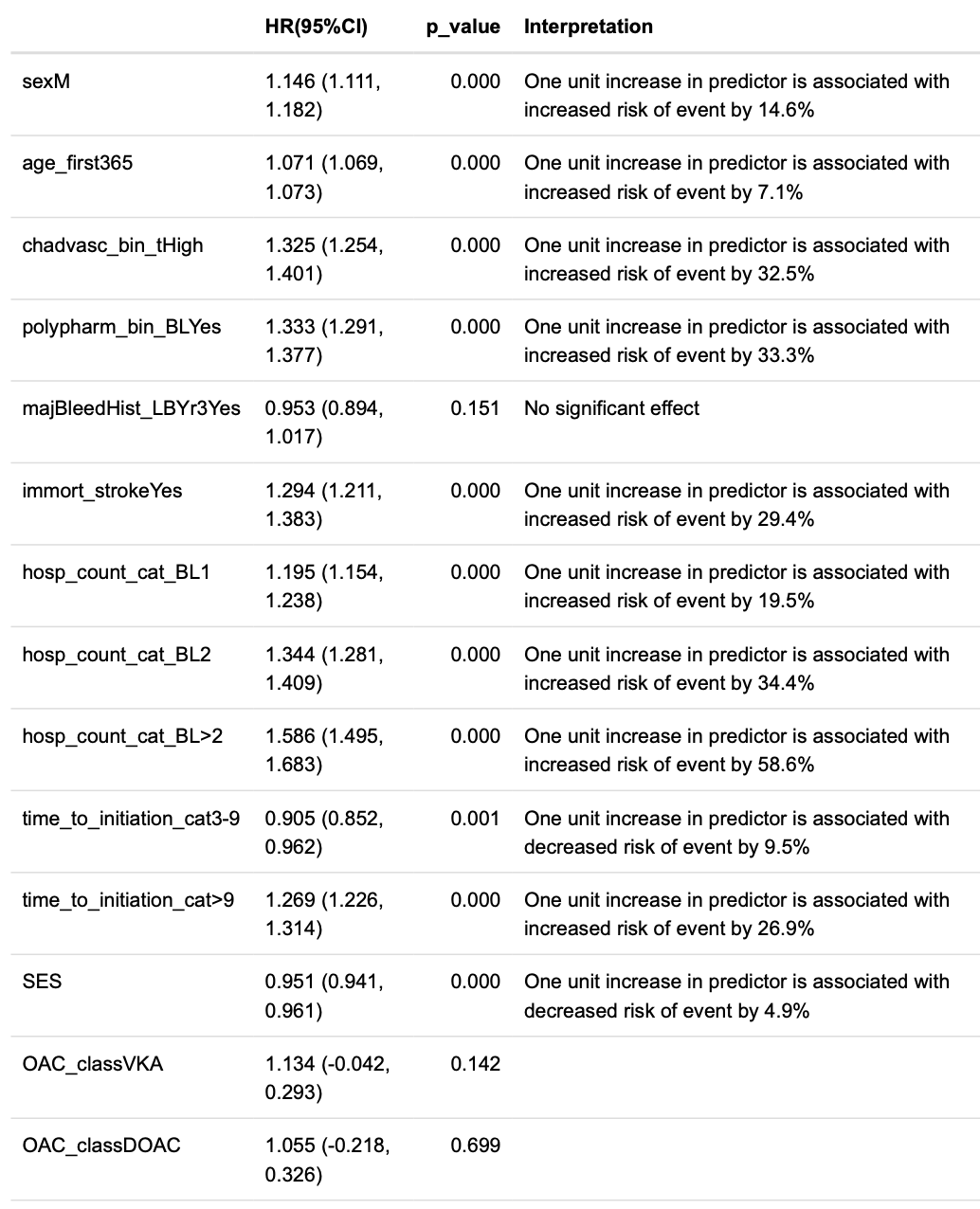

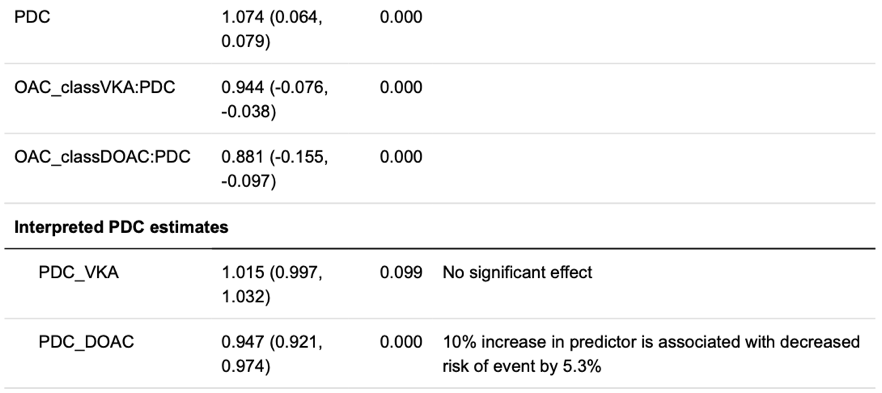

##### Table A6.4: Cox proportional hazards model of time to SSE

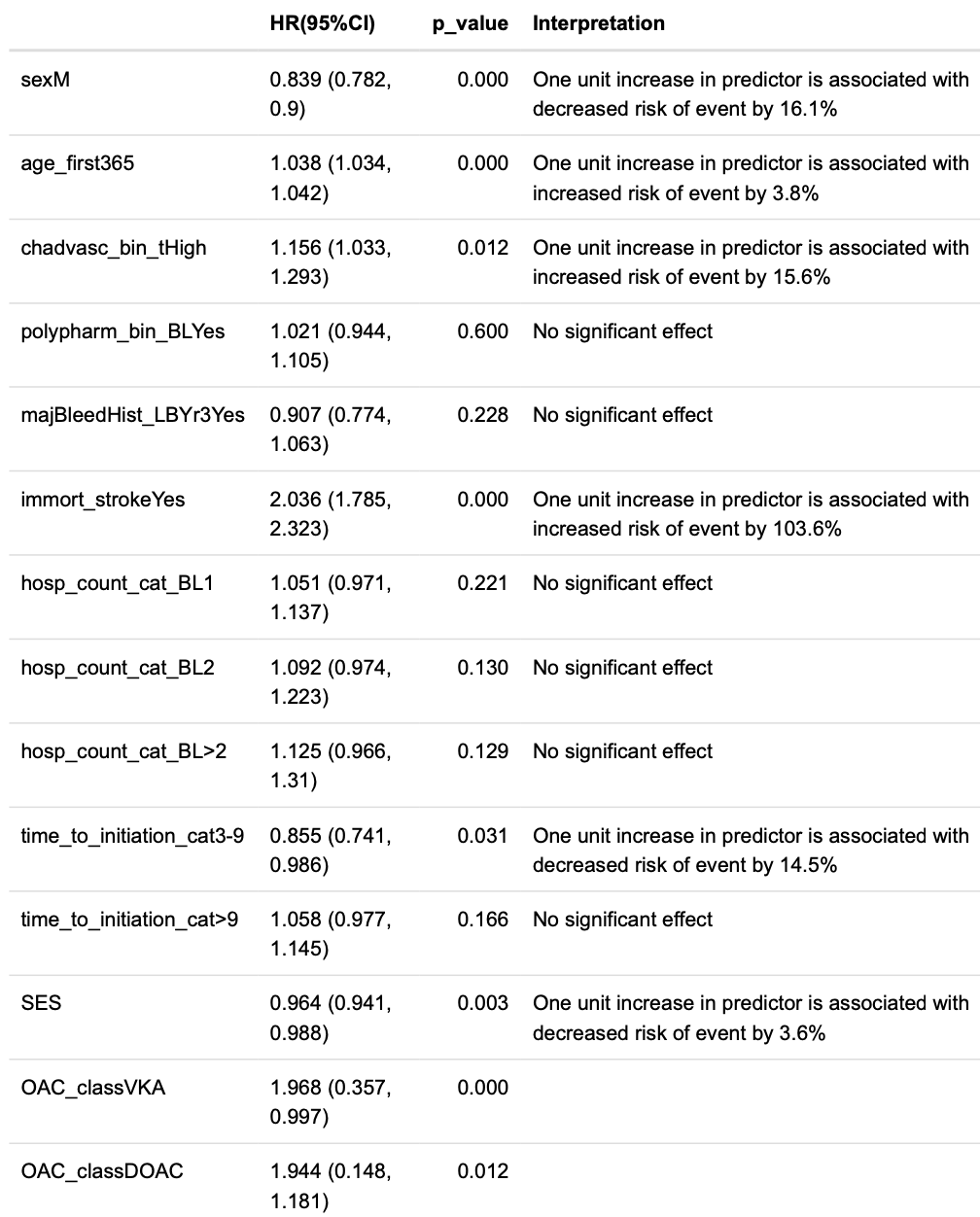

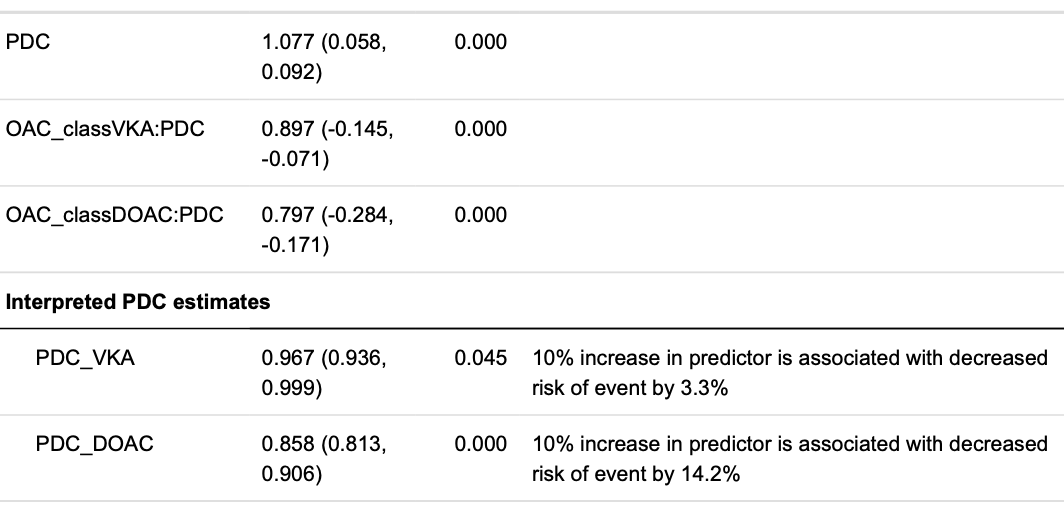

##### Table A6.5: Cox proportional hazards model of time to SSE or TIA

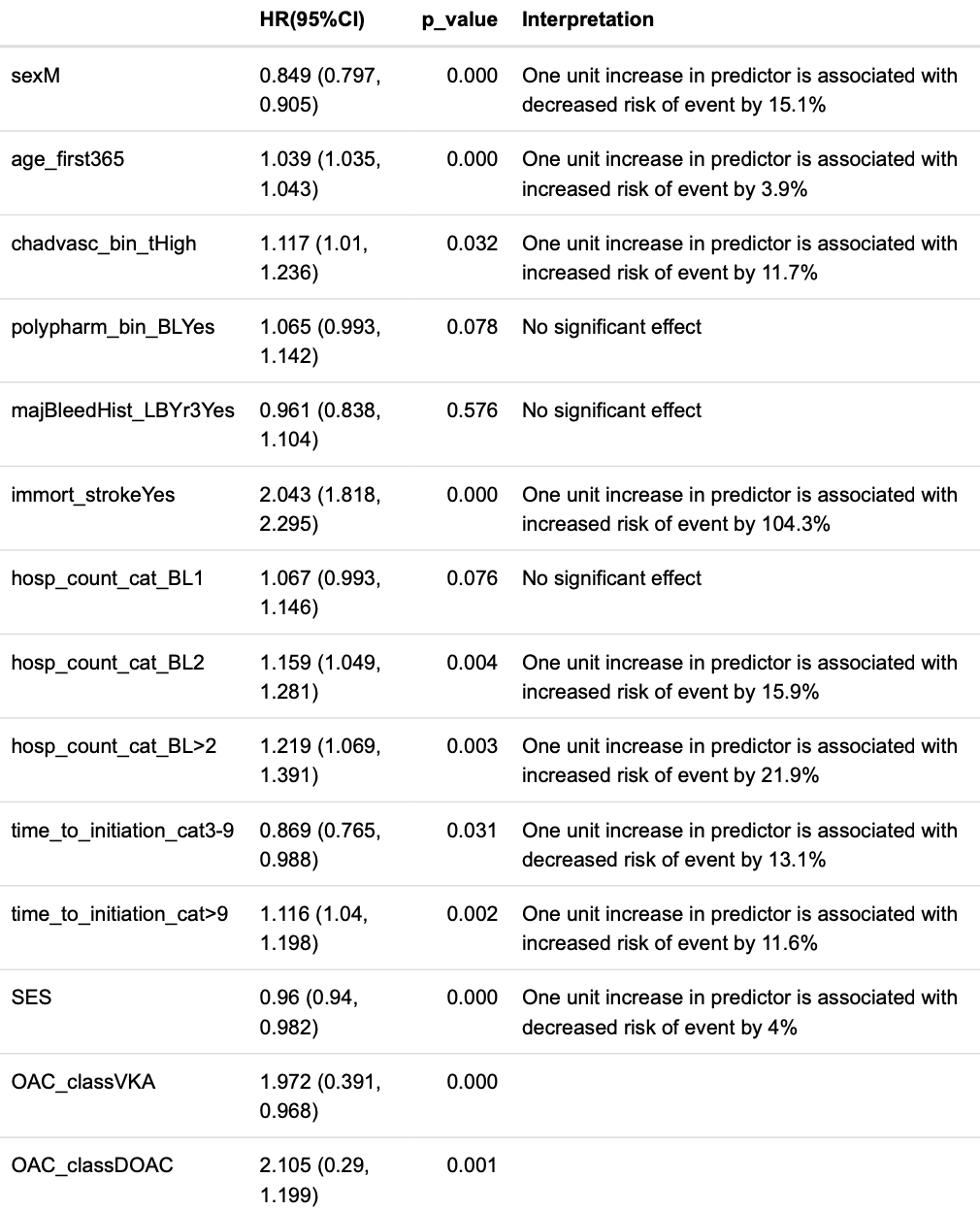

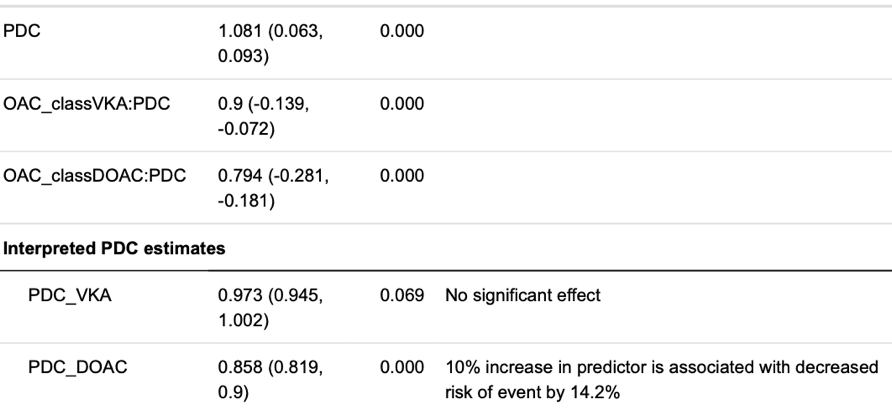

##### Table A6.6: Cox proportional hazards model of time to ischemic stroke

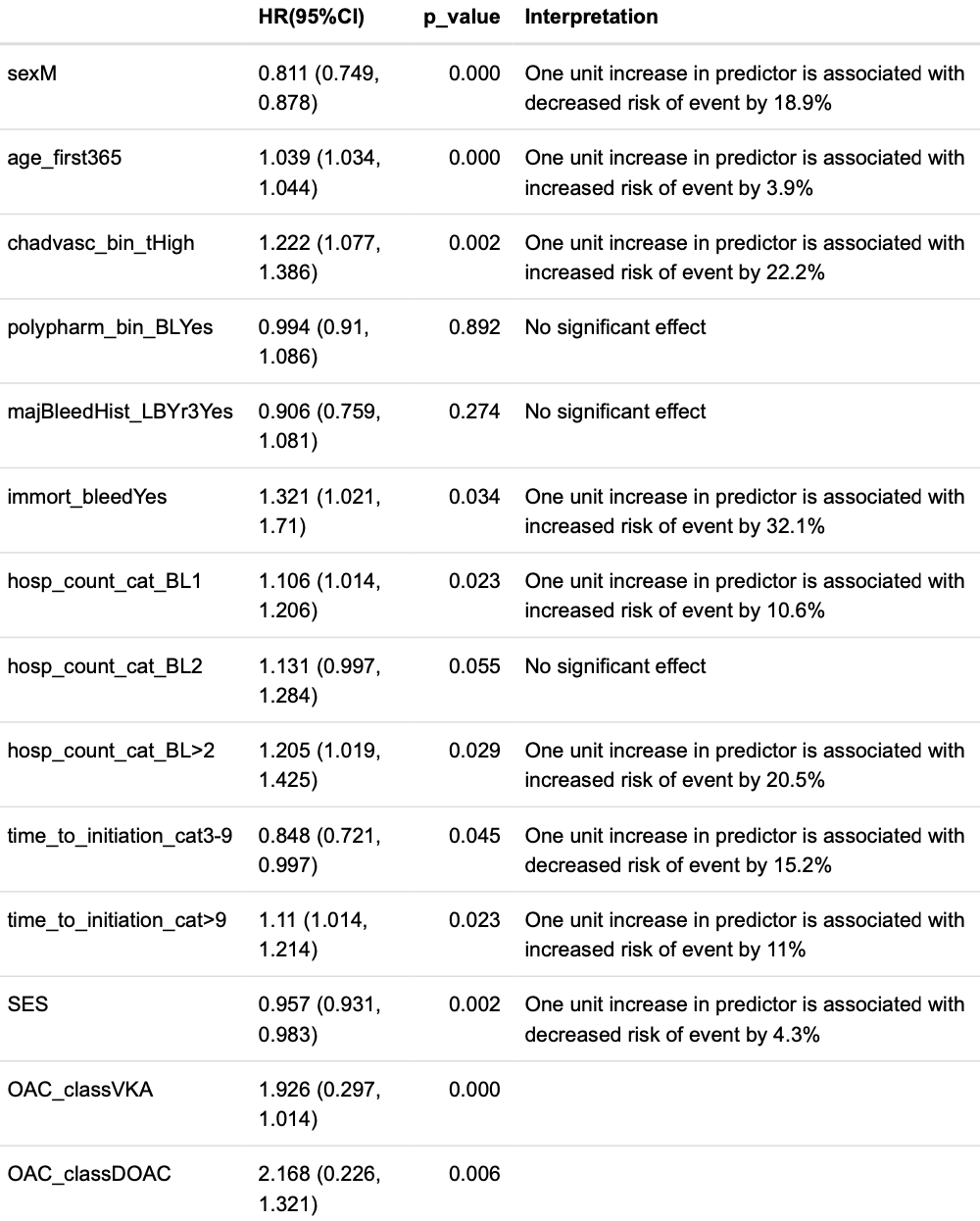

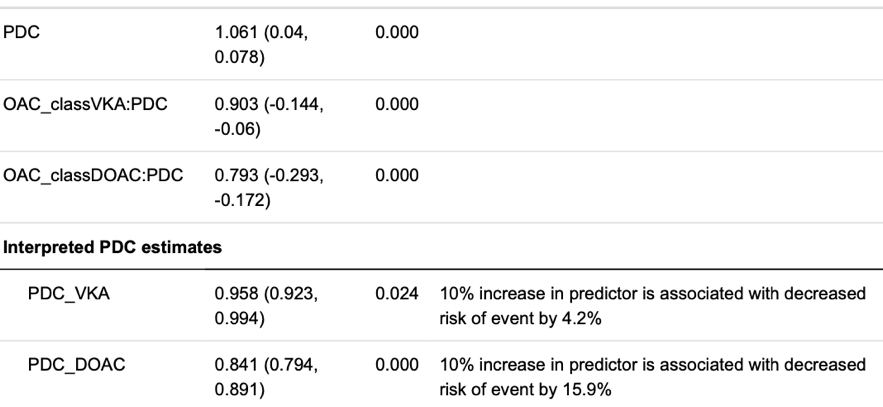

##### Table A6.7: Cox proportional hazards model of time to death

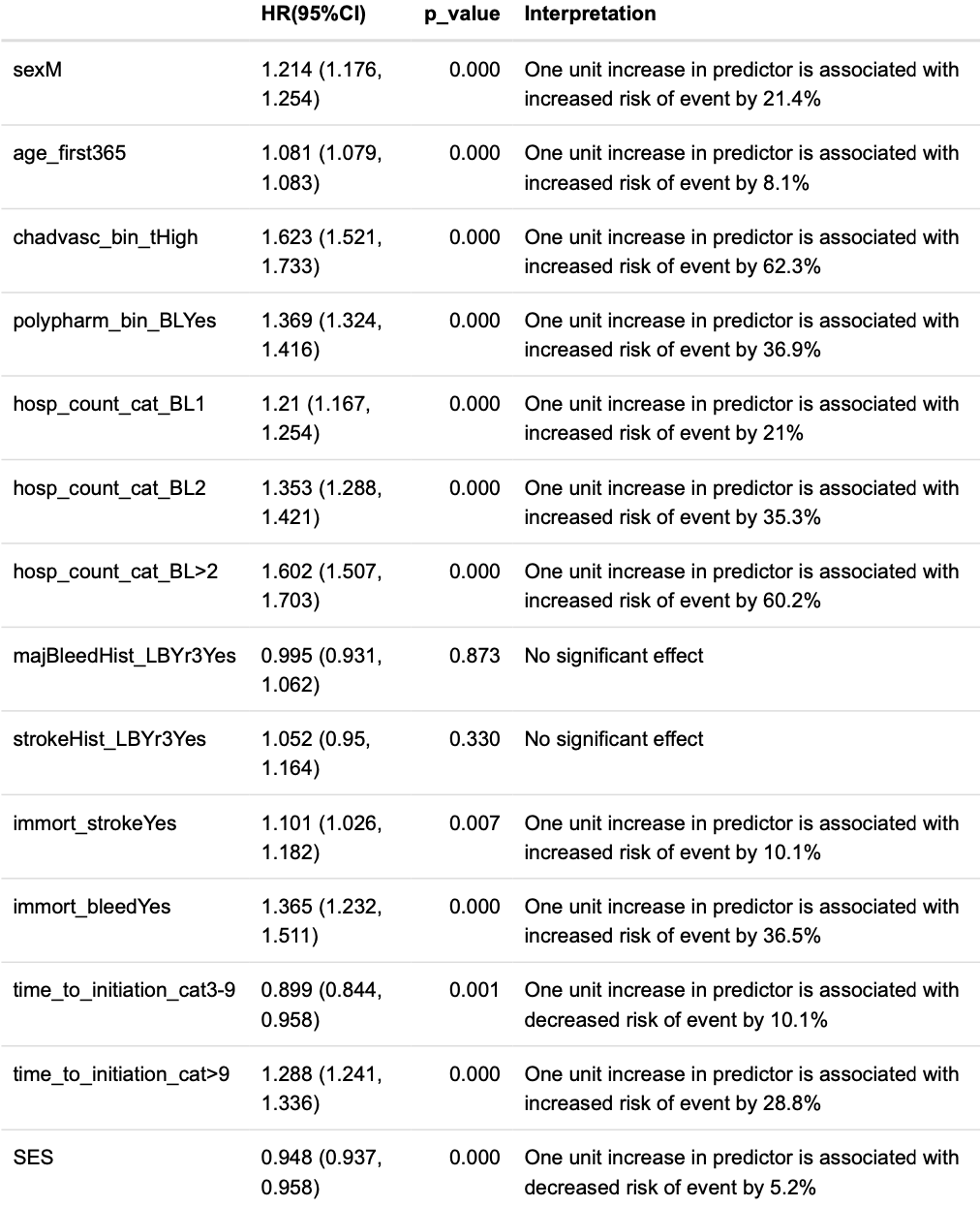

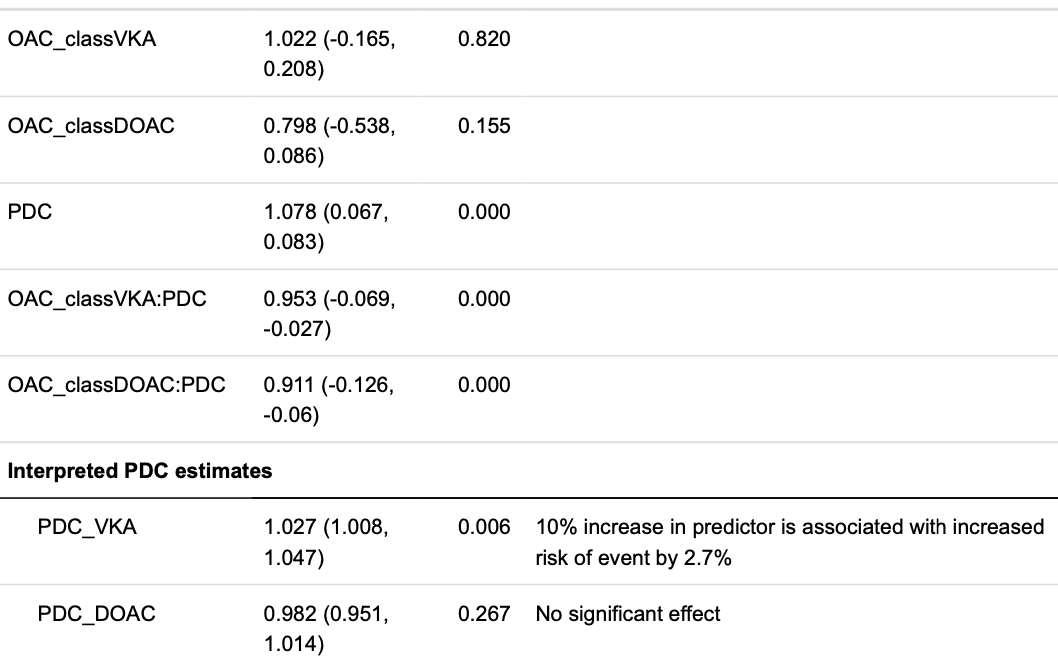

##### Table A6.8: Cox proportional hazards model of time to cardiovascular death

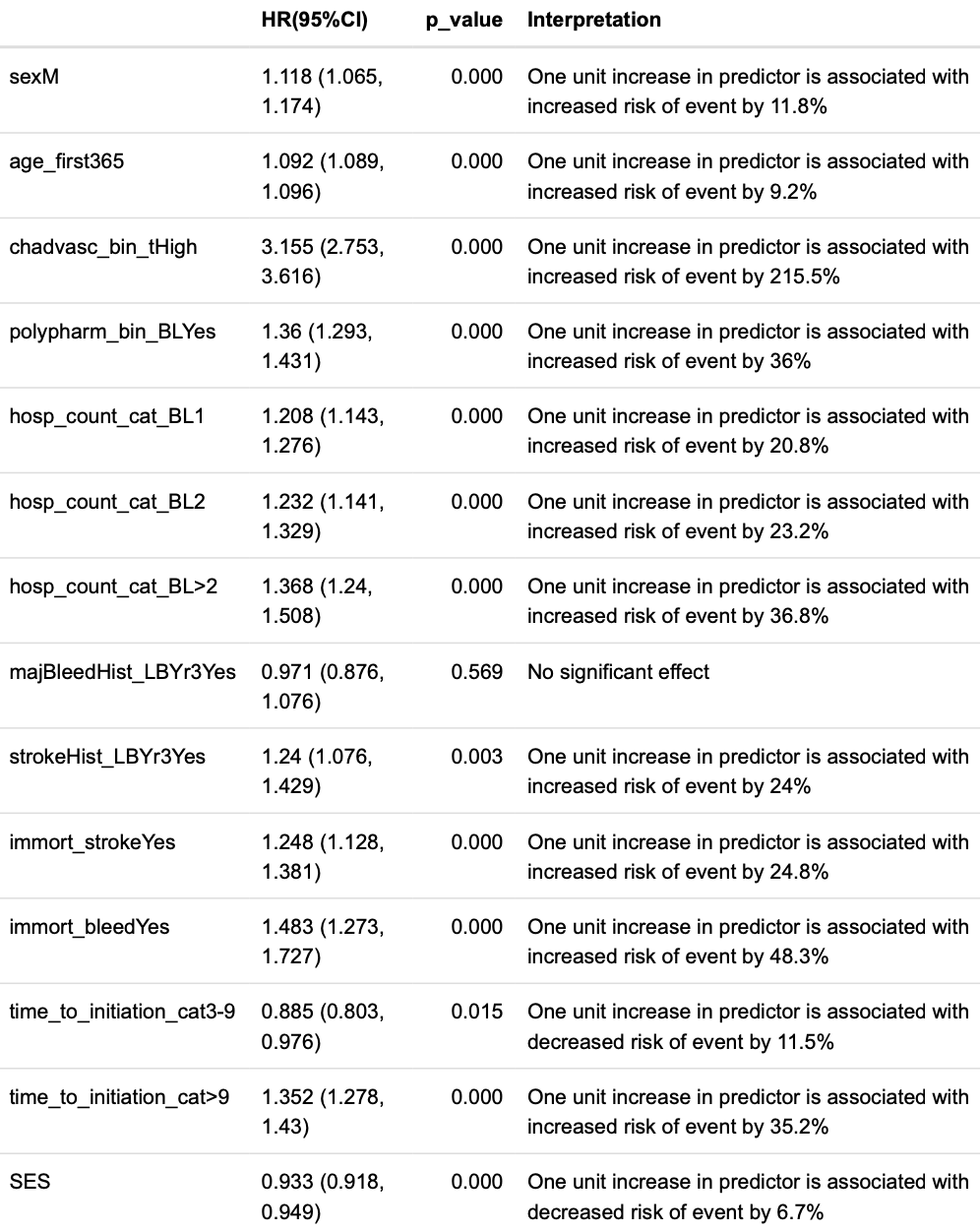

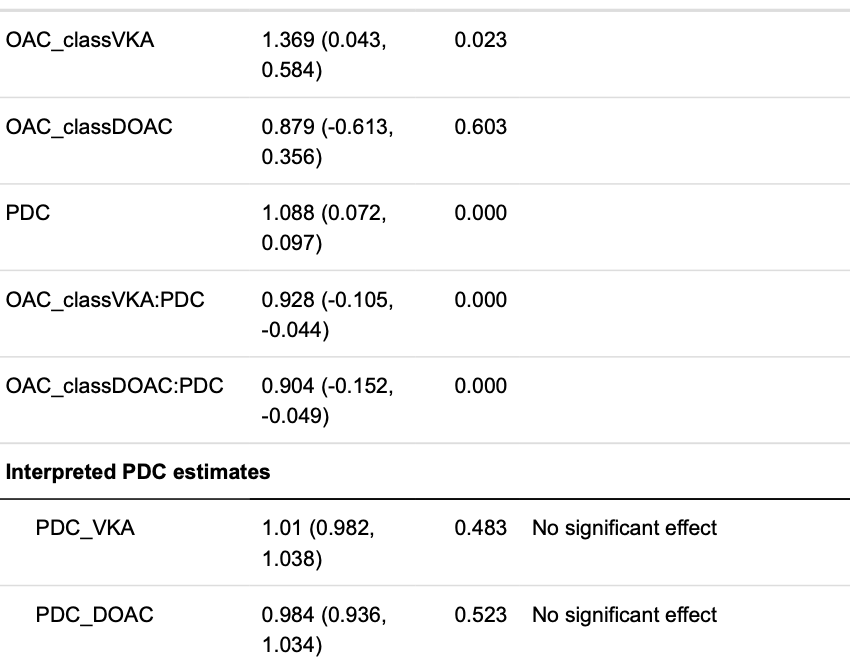

##### Table A6.9: Cox proportional hazards model of time to first major bleeding event

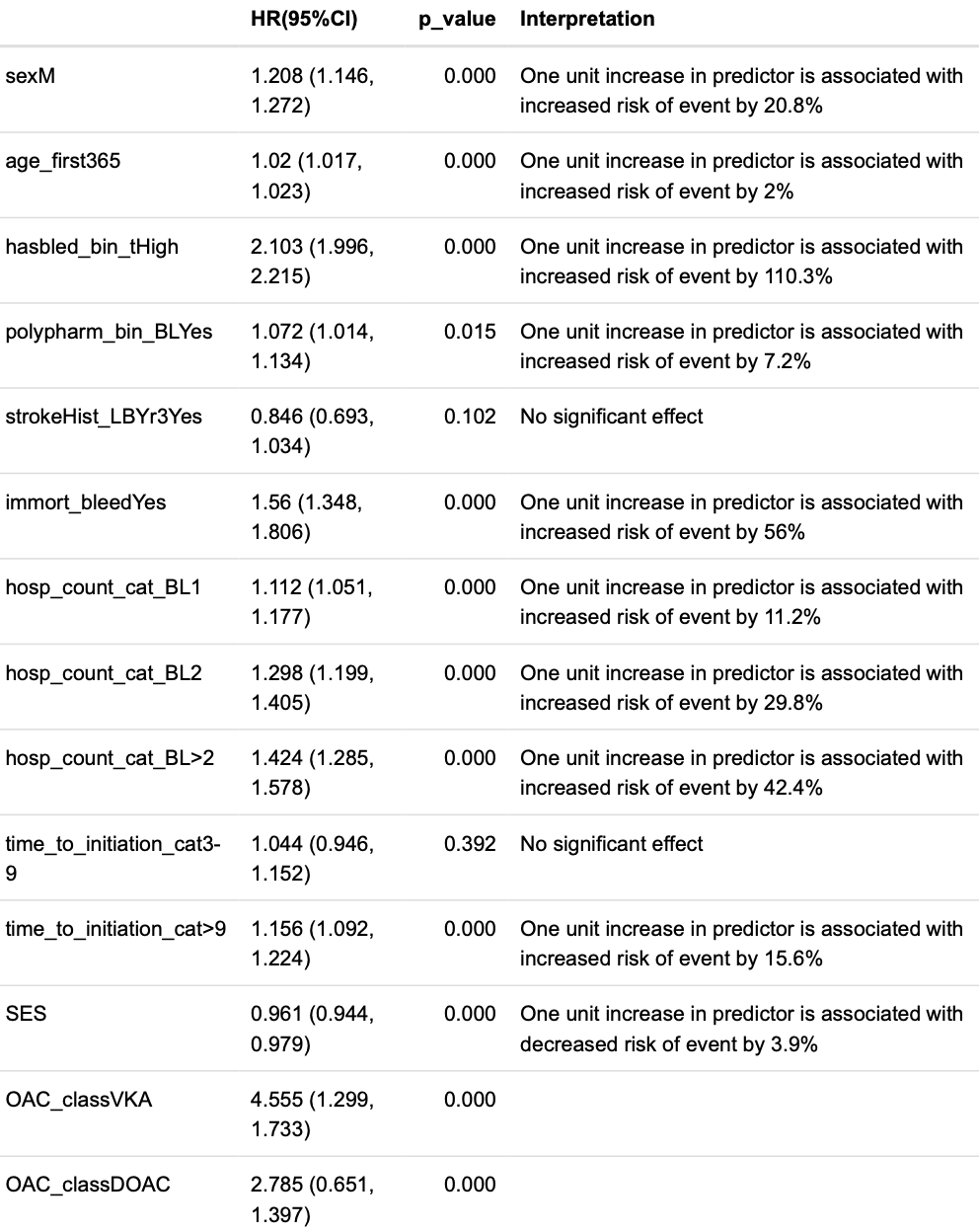

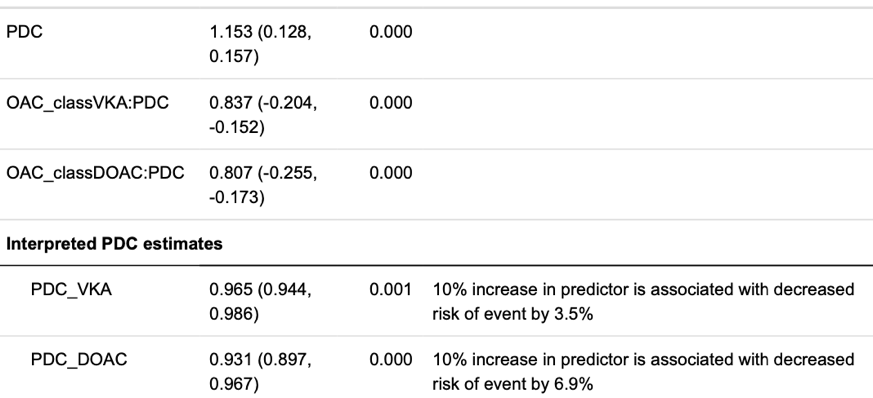

##### Table A6.10: Sex-specific Cox proportional hazards models

**Females**

*stroke, TIA, or death*

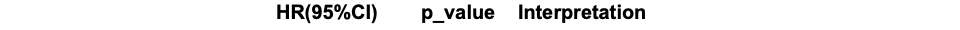

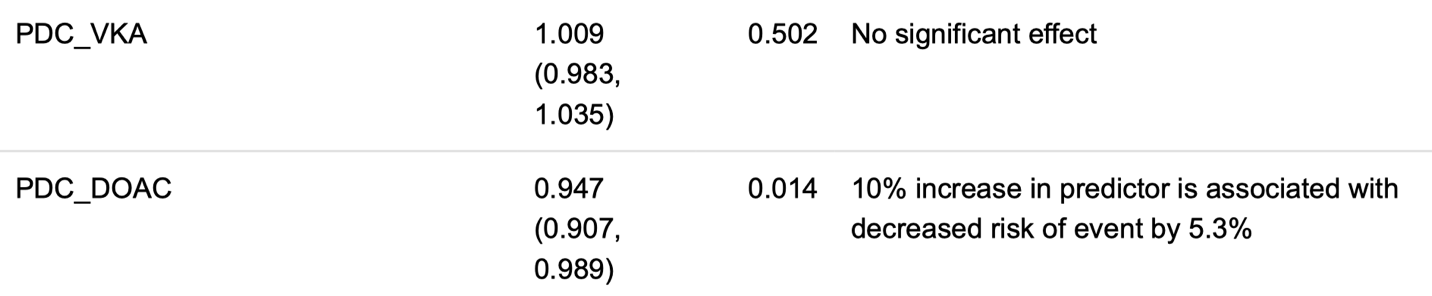

*SSE*

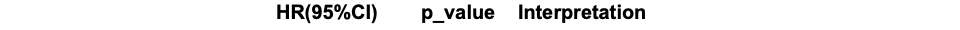

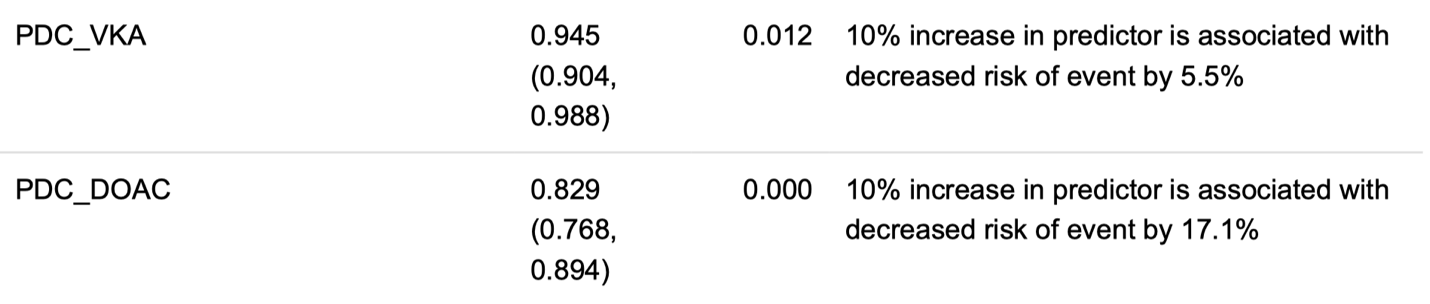

**Males**

*stroke, TIA, or death*

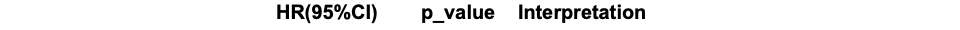

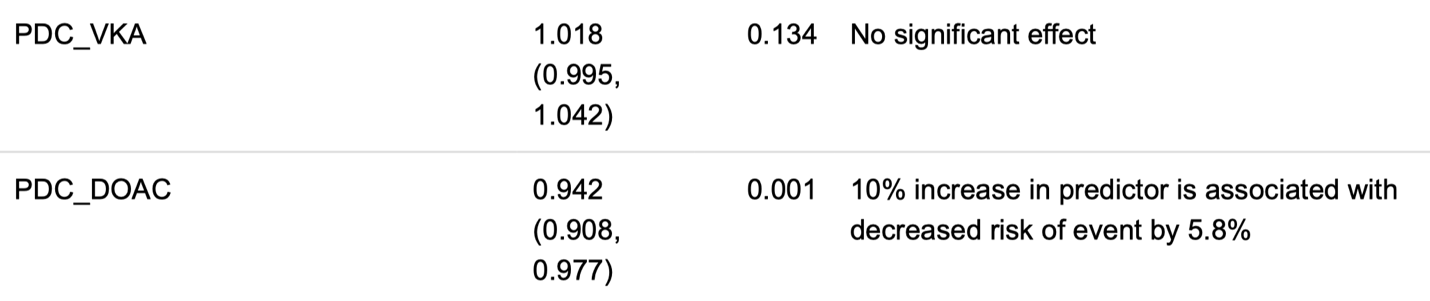

*SSE*

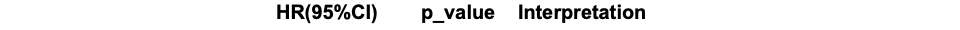

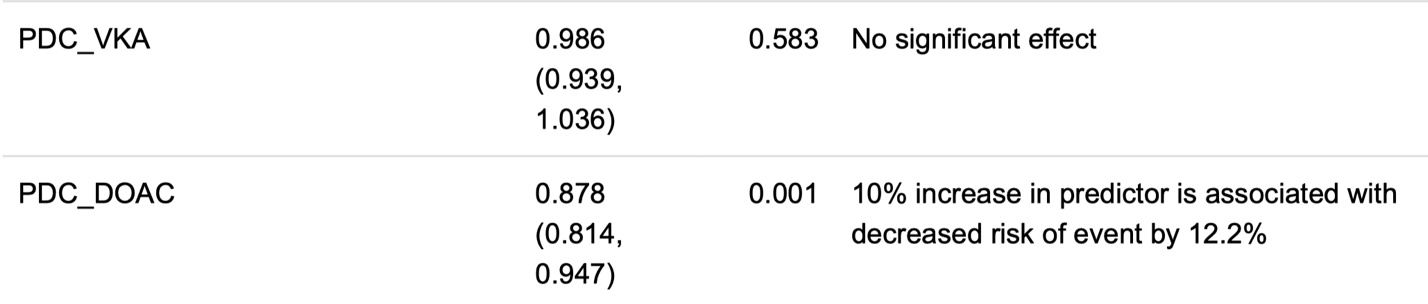

Table A7: Frequency of Outcomes Stratified by OAC Class and individual OAC

##### Table A7.1: Frequency of primary outcomes stratified by OAC drug and class

| Event | DOAC | | | | | | | VKA | No OAC at time of event |
| --- | --- | --- | --- | --- | --- | --- | --- | --- | --- |
|  | Apixaban | Dabigatran | | Edoxaban | | Rivaroxaban | |  |  |
| Death (all cause) or Stroke (SSE or TIA) | 338 (14.2%) | | | | | | | 927 (39.0%) | 1112 (46.8%) |
|  | 93 (3.9%) | 97 (4.1%) | | 0 (0.00%) | | 148 (6.2%) | |  |  |
| SSE | 238 (13.2%) | | | | | | | 683 (37.9%) | 883 (48.9%) |
|  | 70 (3.9%) | | 61 (3.4%) | | 0 (0%) | | 107 (5.9%) |  |  |

##### Table A7.2: Frequency of stroke types stratified by OAC drug and class

| Event | DOAC | | | | | | | VKA | No OAC at time of event |
| --- | --- | --- | --- | --- | --- | --- | --- | --- | --- |
|  | Apixaban | Dabigatran | | Edoxaban | | Rivaroxaban | |  |  |
| SSE or TIA | 338 (14.2%) | | | | | | | 927 (39.0%) | 1112 (46.8%) |
|  | 93 (3.9%) | 97 (4.1%) | | 0 (0%) | | 148 (6.2%) | |  |  |
| Ischemic | 200 (13.6%) | | | | | | | 527 (35.9%) | 740 (50.4%) |
|  | 61 (4.2%) | | 53 (3.6%) | | 0 (0%) | | 86 (5.9%) |  |  |
| Hemorrhagic | 27 (15.1%) | | | | | | | 88 (49.2%) | 64 (35.8%) |
|  | 8 (4.5%) | | 4 (2.2%) | | 0 (0%) | | 15 (8.4%) |  |  |

##### Table A7.3: Frequency of major bleeding stratified by OAC drug and class

| Event | DOAC | | | | | | | VKA | No OAC at time of event |
| --- | --- | --- | --- | --- | --- | --- | --- | --- | --- |
|  | Apixaban | Dabigatran | | Edoxaban | | Rivaroxaban | |  |  |
| Major bleeding | 837 (21.8%) | | | | | | | 1904 (49.6%) | 1098 (28.6%) |
|  | 199 (5.2%) | 210 (5.5%) | | 0 (0%) | | 428 (11.1%) | |  |  |
| Nontraumatic intracranial hemorrhage | 38 (12.7%) | | | | | | | 164 (54.7%) | 98 (32.7%) |
|  | 12 (4.0%) | | 5 (1.7%) | | 0 (0%) | | 21 (7.0%) |  |  |

##### Table A7.4: Frequency of death Stratified by OAC drug and class

| Event | DOAC | | | | | | | VKA | No OAC at time of event |
| --- | --- | --- | --- | --- | --- | --- | --- | --- | --- |
|  | Apixaban | Dabigatran | | Edoxaban | | Rivaroxaban | |  |  |
| All cause | 2348 (17.9%) | | | | | | | 4044 (30.9%) | 6704 (51.2%) |
|  | 841 (6.4%) | 514 (3.9%) | | 1 (0.00%) | | 992 (7.6%) | |  |  |
| CVS | 936 (17.6%) | | | | | | | 1750 (33.0%) | 2620 (49.4%) |
|  | 356 (6.7%) | | 191 (3.6%) | | 1 (0.00%) | | 388 (7.3%) |  |  |
| AF | 153 (25.4%) | | | | | | | 145 (24.0%) | 305 (50.6%) |
|  | 64 (10.6%) | | 29 (4.8%) | | 0 (0%) | | 60 (10.0%) |  |  |
| Stroke | 120 (14.6%) | | | | | | | 273 (33.2%) | 430 (52.2%) |
|  | 34 (4.1%) | | 33 (4.0%) | | 0 (0%) | | 53 (6.4%) |  |  |
| Bleed | 56 (18.4%) | | | | | | | 148 (48.5%) | 101 (33.1%) |
|  | 20 (6.6%) | | 10 (3.3%) | | 0 (0%) | | 26 (8.5%) |  |  |

#### Table A8: Characteristics of the study cohort and patients who were excluded due to filling only one OAC prescription vs. included patients

Study method: “…because ≥2 OAC prescription fills were required to measure adherence, patients with only one OAC prescription fill were excluded.”

| **Patient characteristic** | **Patients with ≥2 OAC prescription fills after index date [study cohort]**  **(n = 34946)** | **Patients with only 1 OAC prescription after index date [excluded]**  **(n = 2136)** |
| --- | --- | --- |
| Female | 15567 (44.5%) | 924 (43.3%) |
| Age, mean (SD) | 70.1 y (11.3) | 68.6 (14.6) |
| Age category  <55  55-65  65-75  ≥75 | 3183 (9.1%)  6983 (20.0%)  11413 (32.7%)  13367 (38.3%) | 367 (17.2%)  383 (17.9%)  543 (25.4%)  843 (39.5%) |
| Neighborhood income quintile | 3.0 ± 1.4 | 3.1 ± 1.6 |
| Socioeconomic status category  <2  2-3  3-4  ≥4 | 7335 (21.0%)  7132 (20.4%)  6862 (19.6%)  13617 (39.0%) | 416 (19.5%)  434 (20.3%)  402 (18.8%)  884 (41.4%) |
| Index OAC  Warfarin  Dabigatran  Rivaroxaban  Apixaban  Edoxaban | 24927 (71.3%)  2601 (7.4%)  4714 (13.5%)  2693 (7.7%)  11 (0.0%) | 1303 (61.0%)  168 (7.9%)  457 (21.4%)  203 (9.5%)  5 (0.2%) |
| **Characteristics measured during baseline period (1 year prior to index date)** | | |
| CHA_2_DS_2_-VASc score, median (IQR) | 3 (2, 4) | 2 (1, 4) |
| HAS-BLED score, median (IQR) | 2 (1, 3) | 2 (1, 3) |
| Weighted Charlson Comorbidity Index, median (IQR) | 5 (3, 8) | 4 (2, 8) |
| Hypertension | 17546 (50.2%) | 1040 (48.7%) |
| Vascular disease | 9723 (27.8%) | 531 (24.9%) |
| Diabetes | 6655 (19.0%) | 379 (17.7%) |
| Congestive heart failure | 5461 (15.6%) | 358 (16.8%) |
| Stroke/TIA | 3427 (9.8%) | 139 (6.5%) |
| Abnormal renal function | 2248 (6.4%) | 190 (8.9%) |
| Major bleeding | 2778 (7.9%) | 212 (9.9%) |
| Abnormal liver function | 353 (1.0%) | 32 (1.5%) |
| Alcohol use | 213 (0.6%) | 22 (1.0%) |
| Prescribed nonsteroidal anti-inflammatory (NSAID) or antiplatelet drug, median (IQR) | 0 (0-1) | 1 (0-2) |

#### A8: Sensitivity analyses

##### 2010+ analysis

SSE, TIA, or death

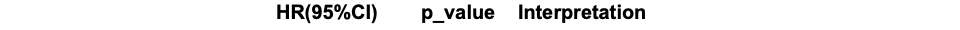

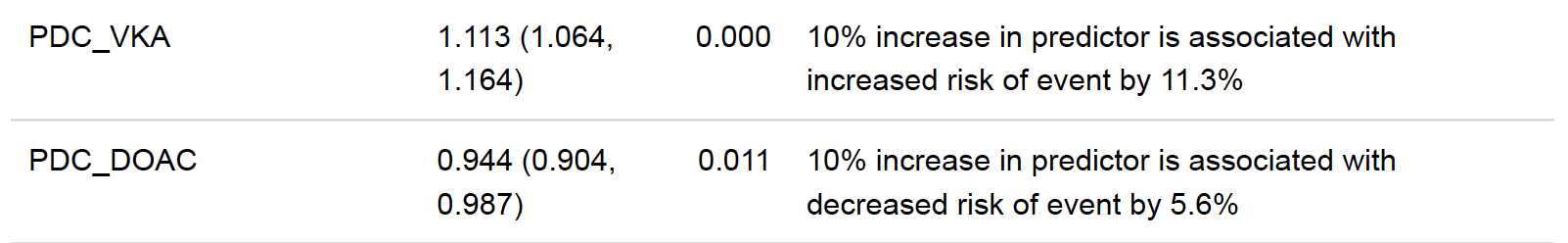

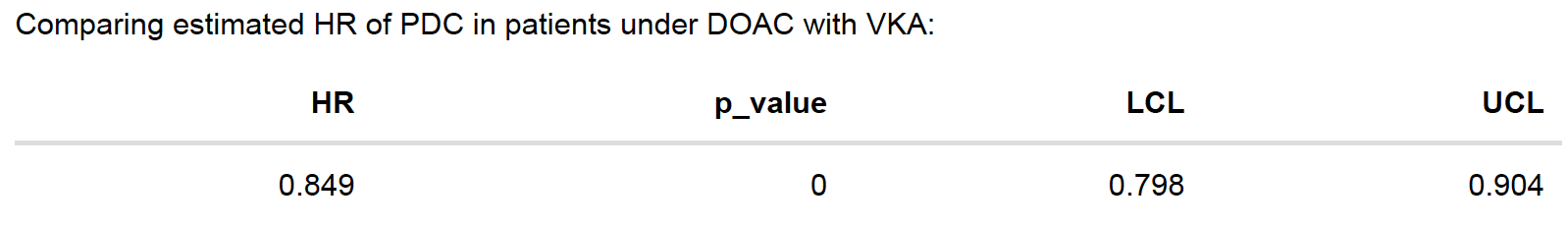

SSE

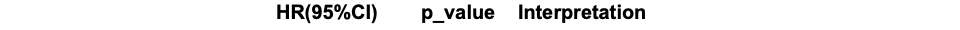

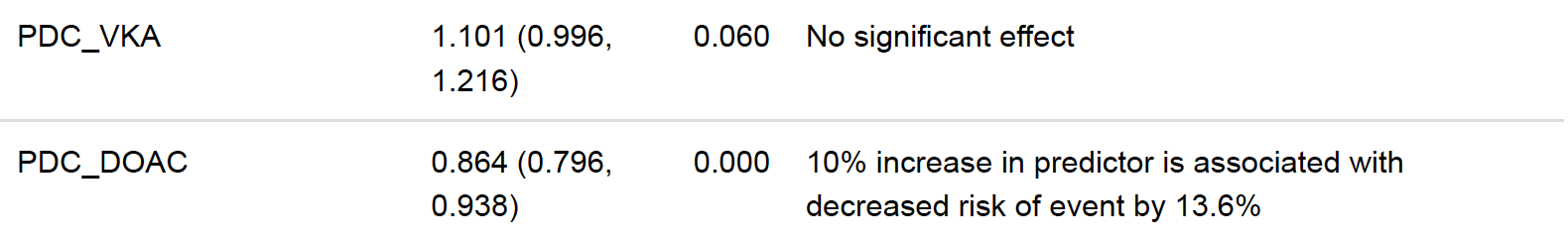

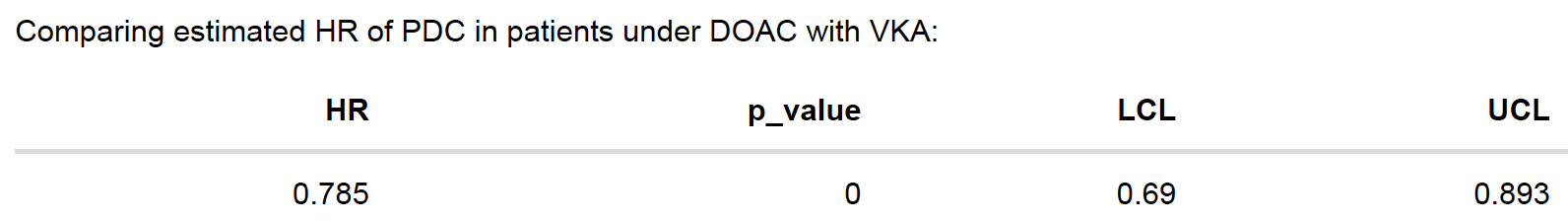

##### varying window of PDC prior to events

**1 month window**

SSE, TIA, or death

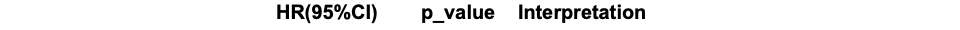

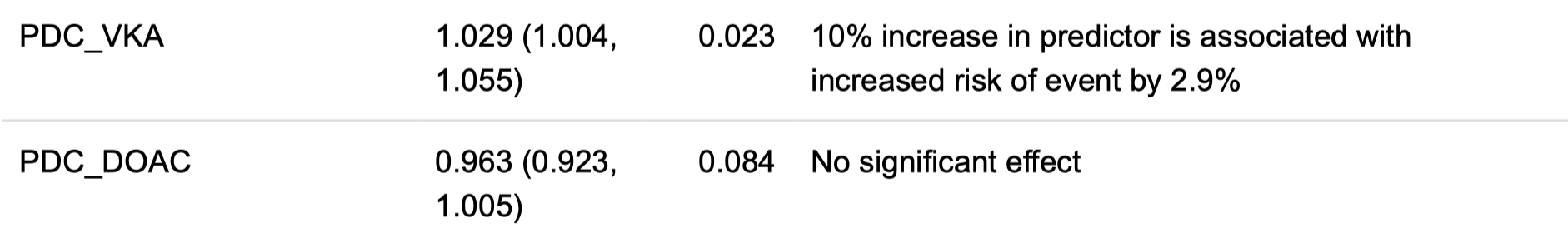

SSE

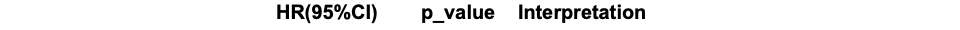

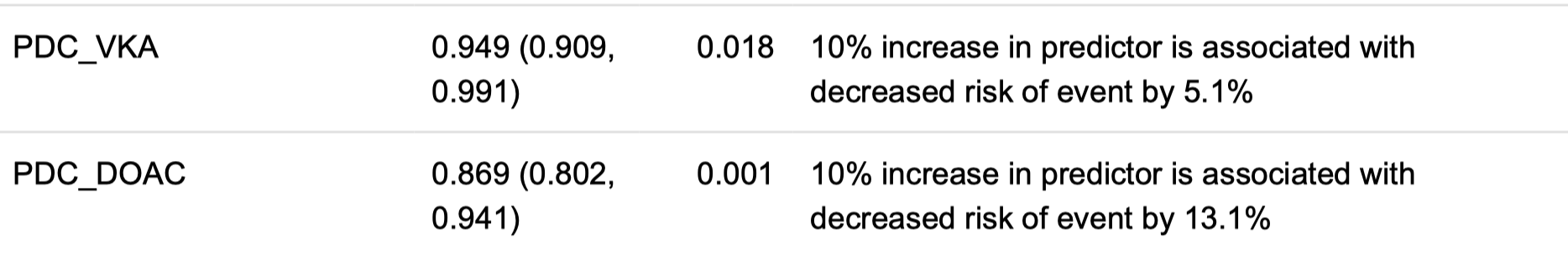

**6-month window**

SSE, TIA, or death

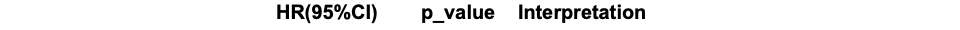

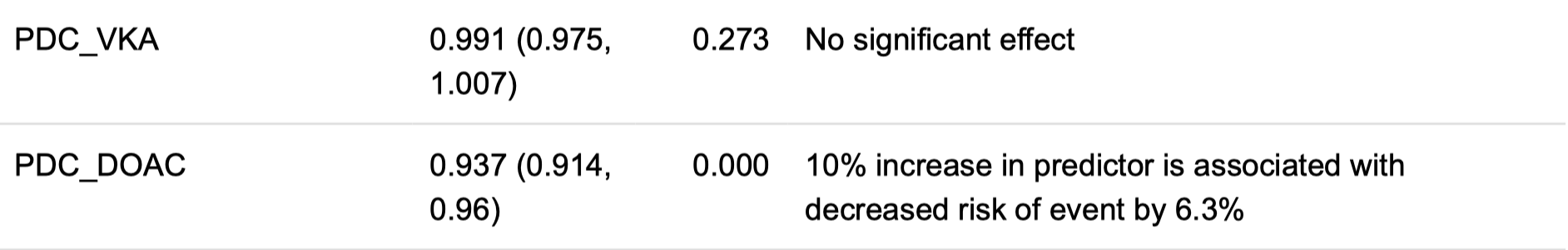

SSE

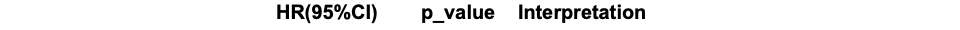

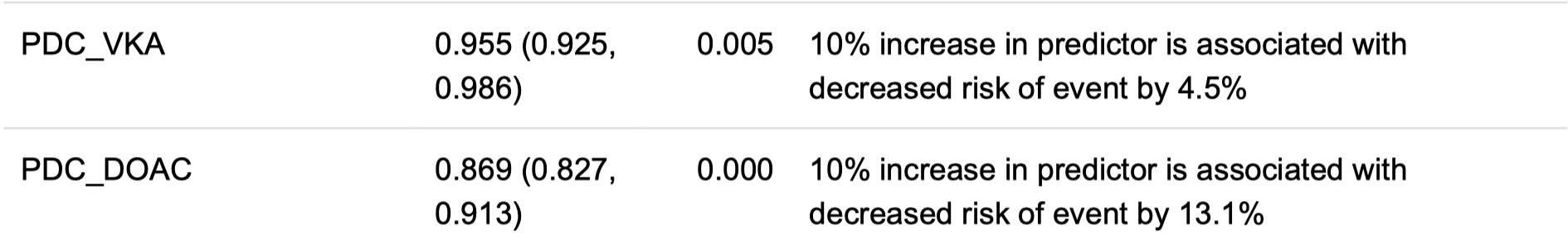

##### robustness of our scheme to attribute drug classes to clinical events

different cut-offs for number of days prior to events for drug changes (5d, 7d, 10d)

**5 days cut-off**

SSE, TIA, or death

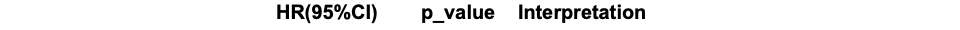

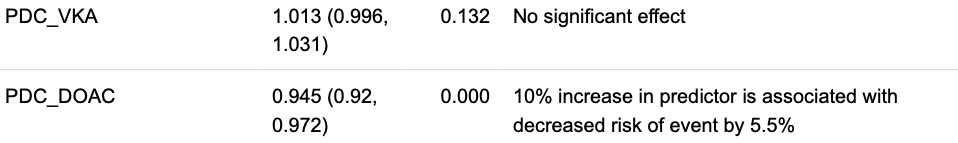

SSE

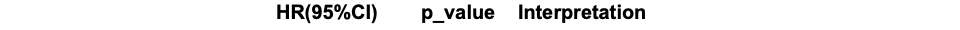

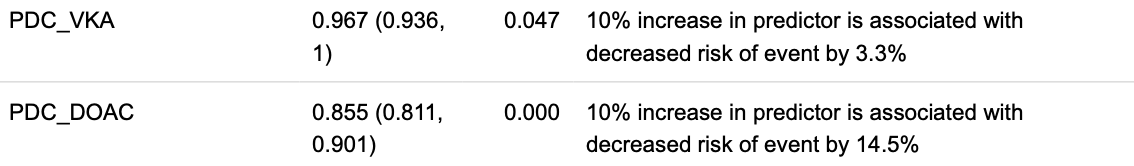

**7 days cut-off**

SSE, TIA, or death

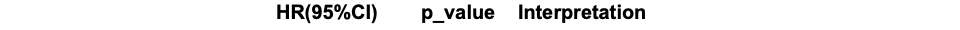

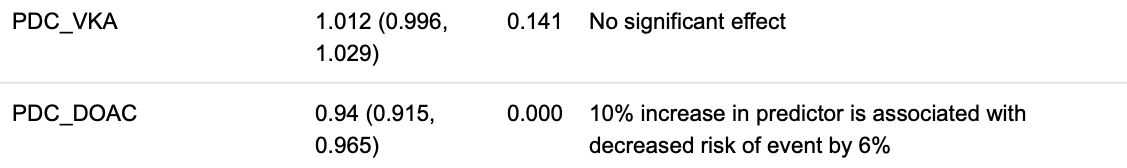

SSE

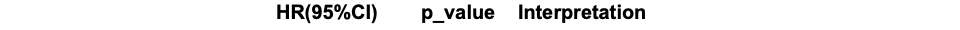

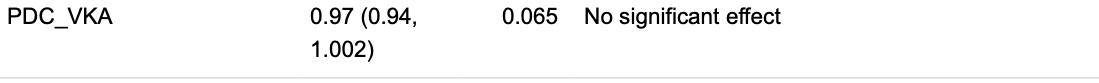

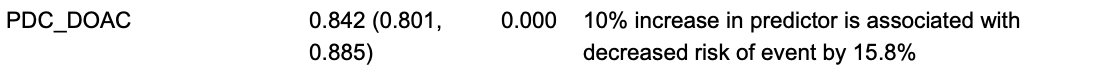

**10 days cut-off**

SSE, TIA, or death

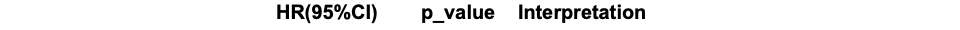

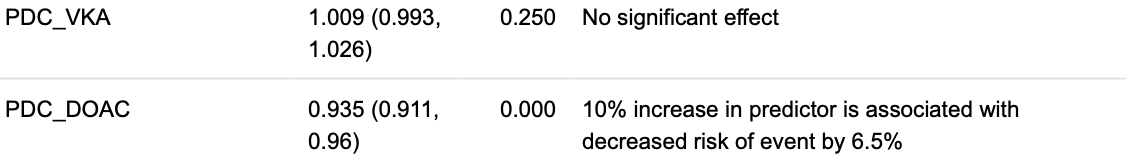

SSE

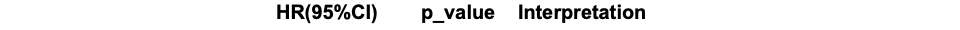

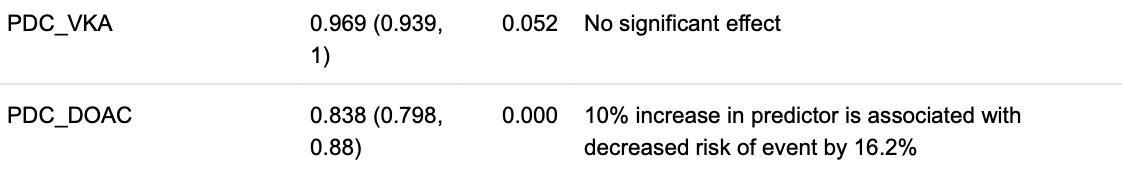

##### excluding patients who had an OAC change within 7 days before of their event (~2000 patients excluded)

SSE, TIA, or death

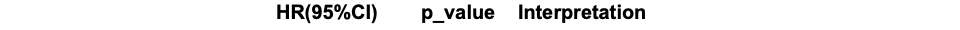

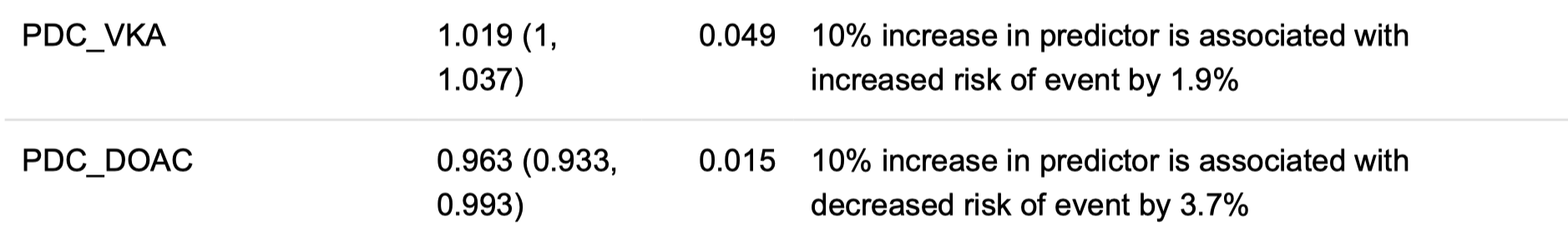

SSE

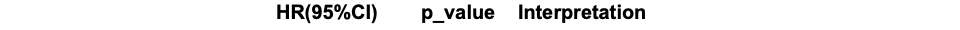

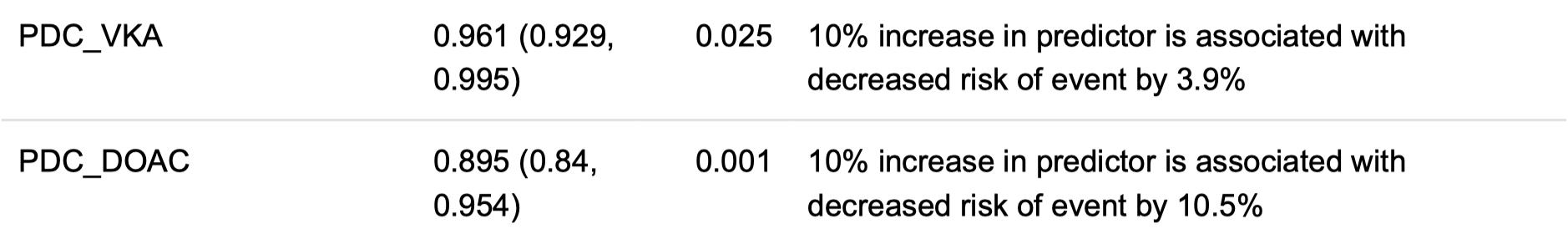

##### excluding patients with a stroke event between diagnosis date and index date (1870 patients excluded)

SSE, TIA, or death

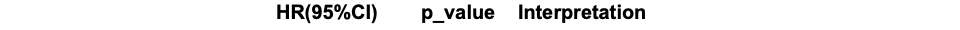

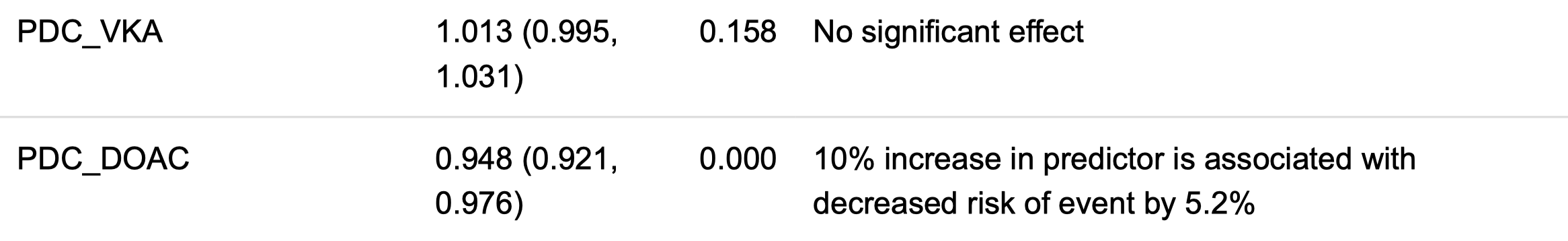

SSE

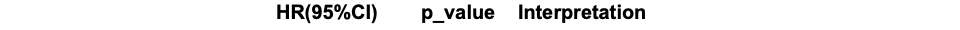

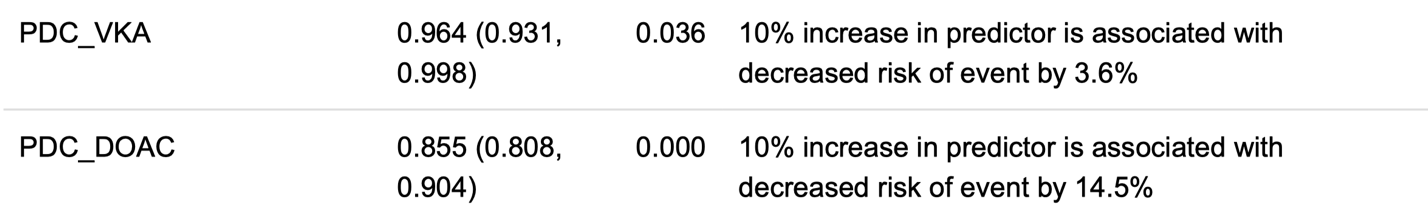

##### falsification (“healthy adherer”) analyses

dental problems as an outcome

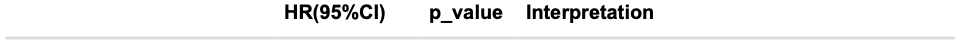

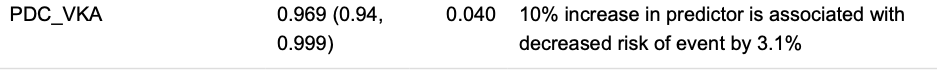

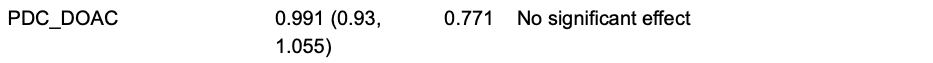

accidental death as an outcome

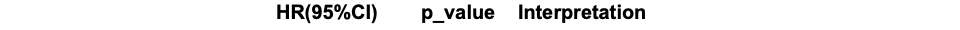

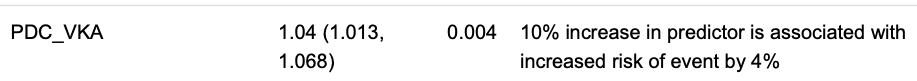

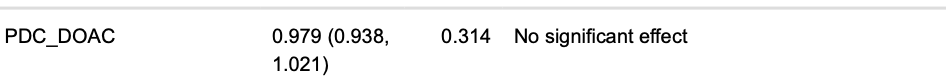
