## Supplementary material for "Association between oral anticoagulant adherence and serious clinical outcomes in patients with atrial fibrillation: A long-term retrospective cohort study": Tables

### Table 1: Characteristics of study patients at index date

| **Patient Characteristic** | **Overall**  **(n =** **44,172)** | **VKA recipients at time of event**  **(n= 12,928) ^i^** | **DOAC recipients at time of event**  **(n=16,688) ^i^** | **Males**  **(n = 24,728) ^i^** | **Females**  **(n = 19,444) ^i^** |
| --- | --- | --- | --- | --- | --- |
| Female | 19,444 (44.0%) |  |  |  |  |
| Age, mean (SD) | 70.6 y (11.4) | 71.7 (10.4) | 69.3 (10.8) | 68.5 (11.7) | 73.4 (10.4) |
| Age category  <55  55-65  65-75  ≥75 | 3865 (8.7%)  8342 (18.9%)  14,115 (32.0%)  17,850 (40.4%) | 793 (6.1%)  2238 (17.3%)  4371 (33.8%)  5526 (42.7%) | 1497 (9.0%)  3796 (22.7%)  5770 (34.6%)  5625 (33.7%) | 2976 (12.0%)  5504 (22.3%)  8073 (32.6%)  8175 (33.1%) | 889 (4.6)  2838 (14.6%)  6042 (31.1%)  9675 (49.8%) |
| Neighborhood income quintile ^d^ | 3.1 ± 1.4 | 3.0 ± 1.4 | 3.2 ± 1.4 | 3.1 ± 1.4 | 3.0 ± 1.4 |
| Socioeconomic status category  <2  2-3  3-4  ≥4 | 8352 (18.9%)  8571 (19.4%)  8770 (19.9%)  18,479 (41.8%) | 2532 (19.6%)  2602 (20.1%)  2667 (20.6%)  5127 (39.7%) | 2884 (17.3%)  3091 (18.5%)  3258 (19.5%)  7455 (44.7%) | 4322 (17.5%)  4540 (18.4%)  4905 (19.8%)  10,961 (44.3%) | 4030 (20.7%)  4031 (20.7%)  3865 (19.9%)  7518 (38.7%) |
| Index OAC  Warfarin  Dabigatran  Rivaroxaban  Apixaban  Edoxaban | 31,744 (71.9%)  3281 (7.4%)  5825 (13.2%)  3301 (7.5%)  21 (0.0%) | 12,370 (28.0%)  242 (0.5%)  235 (0.5%)  81 (0.2%)  0 (0.0%) | 8153 (15.5%)  2079 (4.7%)  4027 (9.1%)  2412 (5.5%)  17 (0.0%) | 17,803 (72.0%)  1843 (7.5%)  3268 (13.2%)  1802 (7.3%)  12 (0.0%) | 13,941 (71.7%)  1438 (7.4%)  2557 (13.2%)  1499 (7.7%)  9 (0.0%) |
| **Characteristics measured during baseline period (1 year prior to index date)** | | | | | |
| CHA_2_DS_2_-VASc score, median (IQR) ^e^ | 3 (2, 4) | 3 (2, 4) | 3 (2, 4) | 2 (1, 4) | 3 (2, 4) |
| HAS-BLED score, median (IQR) ^f^ | 2 (1, 3) | 2 (2, 3) | 2 (1, 3) | 2 (1, 3) | 2 (2, 3) |
| Charlson Comorbidity Index, median (IQR) ^g^ | 4 (3, 5) | 4 (3, 5) | 3 (2, 4) | 3 (2, 4) | 4 (3, 5) |
| Elixhauser Comorbidity Index, median (IQR)^g^ | 5 (3, 10) | 5 (3, 10) | 5 (3, 9) | 5 (3, 9) | 5 (3, 10) |
| Hypertension | 26,371 (59.7%) | 7,853 (60.7%) | 10,169 (60.9%) | 13,652 (55.2%) | 12,719 (65.4%) |
| Vascular disease | 12,131 (27.5%) | 663 (28.3%) | 4425 (26.5%) | 7262 (29.4%) | 4869 (25.0%) |
| Diabetes | 8699 (19.7%) | 2604 (20.1%) | 3311 (19.8%) | 5274 (21.2%) | 3425 (17.6%) |
| Congestive heart failure | 6881 (15.6%) | 2264 (17.5%) | 2149 (12.9%) | 3808 (15.4%) | 3073 (15.8%) |
| Stroke/TIA history | 4276 (9.7%) | 1326 (10.3%) | 1536 (9.2%) | 2148 (8.7%) | 2128 (10.9%) |
| Abnormal renal function | 2921 (6.6%) | 937 (7.2%) | 956 (5.7%) | 1526 (6.2%) | 1395 (7.2%) |
| Major bleeding history | 3598 (8.1%) | 940 (7.3%) | 1441 (8.6%) | 2199 (8.9%) | 1399 (7.2%) |
| Abnormal liver function | 455 (1.0%) | 101 (0.8%) | 192 (1.2%) | 277 (1.1%) | 178 (0.9%) |
| Alcohol use | 252 (0.6%) | 45 (0.3%) | 95 (0.6%) | 204 (0.8%) | 48 (0.2%) |
| Prescribed nonsteroidal anti-inflammatory (NSAID) or antiplatelet drug, median (IQR)^h^ | 0 (0-1) | 0 (0-1) | 0 (0-1) | 0 (0-1) | 0 (0-1) |
| **SSE and major bleeding risk scores measured during event period (1 year prior to event date or end of follow-up)** | | | | | |
| CHA_2_DS_2_-VASc score, median (IQR) ^e^ | 4 (3, 5) | 4 (3, 5) | 3 (2, 4) | 4 (2, 5) | 4 (3, 5) |
| HAS-BLED score, median (IQR) ^f^ | 2 (2, 3) | 2 (2, 3) | 2 (2, 3) | 2 (2, 3) | 2 (2, 3) |
| Charlson Comorbidity Index, median (IQR)^g^ | 4 (3, 5) | 5 (4, 6) | 4 (3, 5) | 4 (2, 5) | 4 (3, 6) |
| Elixhauser Comorbidity Index, median (IQR)^g^ | 5 (0, 12) | 8 (3, 17) | 3 (0, 7) | 5 (0, 12) | 5 (0, 12) |

OAC: Oral Anticoagulants. TIA: Transient ischemic attack. IQR: Interquartile range.

Values are n (%), mean ± standard deviation, median (IQR), unless otherwise stated.

^d^ Neighborhood income quintile represents the average equivalized disposable income by postal code in patients’ residential area at index date, with 1 as the lowest and 5 as the highest level.

^e^ Stroke risk score calculated based on the presence of comorbidities: Cardiomyopathy (1 point), Hypertension (1 point), Age≥75 (2 points), Diabetes (1 point), Stroke (2 points), Vascular disease (1 point), Age 65–75 (1 point), Sex category female (1 point). Scored if at least one disease specific ICD code was present in either outpatient or inpatient records during the baseline period. Ascertainment scheme is in Supplemental appendix Table A1.

^f^ Bleeding risk score calculated based on the presence of comorbidities: Hypertension (1 point), Abnormal liver/kidney function (1 point), Stroke (1 point), Bleeding (1 point), Age > 75 years (1 point), Drugs (NSAID or antiplatelet) or alcohol use (1 point). Scored if at least one disease specific ICD code was present in either outpatient or inpatient records during the baseline period. Ascertainment scheme is in Supplemental appendix Tables A1 and A2.

^g^ Each comorbidity category has an associated weight (from 1 to 6), based on the adjusted risk of mortality or resource use, and the sum of all the weights results in a single comorbidity score for a patient. Possible range of scores: 0-31. Zero indicates that no comorbidities were found. Ascertainment scheme is in Supplemental appendix Table A1.

^h^ NSAIDs and ASA could be obtained with or without a prescription in the study jurisdiction.

^i^ Percentages are calculated within each group (OAC class or Sex).

### Table 2: Study outcomes

| **Outcome model** | **Frequency** | **Median time to event in years (IQR)** |
| --- | --- | --- |
| **Primary outcomes** |  |  |
| SSE, TIA, or death | 17,052 | 5.3 (2.6-9.1) |
| SSE | 3257 | 4.1 (1.7-7.6) |
| **Secondary outcomes** |  |  |
| SSE or TIA | 4051 | 4.0 (1.6-7.5) |
| Ischemic stroke | 2588 | 4.1 (1.7-7.5) |
| Death (any cause) | 15,785 | 5.9 (3.1-9.7) |
| Cardiovascular death | 6766 | 6.1 (3.2-9.9) |
| Major bleeding | 6190 | 3.6 (1.4-6.9) |
| Nontraumatic intracranial hemorrhage | 627 | 3.9 (1.6-7.9) |

SSE: stroke or systemic embolism. Ascertainment scheme in Supplemental appendix.
TIA: transient ischemic attack. Ascertainment scheme in Supplemental appendix.
Study outcome definitions are in Supplemental appendix Table A3.

### Table 3: Effects of OAC adherence on clinical outcomes by OAC class

| **Outcome** | **Number of events** | **Drug** | **HR (95% CI)*** | **Change in hazard of outcome per 10% absolute decrease in PDC (95%CI)** | **Contrast test for DOAC vs. VKA**  **HR (95%CI); p-value** | **Detailed model outputs** |
| --- | --- | --- | --- | --- | --- | --- |
| SSE, TIA, or death  (co-primary outcome) | 17,052 | VKA | 1.015 (0.997 – 1.032) | NS | 0.933 (0.903-0.964); p<0.001 | Supplemental materials A 6.3 |
|  |  | DOAC | 0.947 (0.921 – 0.974) | 5.3% (2.6-7.9) increase |  |  |
| SSE  (co-primary outcome) | 3257 | VKA | 0.967 (0.936 – 0.999) | 3.3% (0.1-6.4) increase | 0.888 (0.833-0.946); p<0.001 | Supplemental materials A 6.4 |
|  |  | DOAC | 0.858 (0.813 – 0.906) | 14.2% (9.4-18.7) increase |  |  |
| SSE or TIA | 4051 | DOAC | 0.858 (0.819 – 0.900) | 14.2% (10.0-18.1) increase | 0.882 (0.834-0.933); p<0.001 | Supplemental materials A 6.5 |
|  |  | VKA | 0.958 (0.923 – 0.994) | 4.2% (0.6-7.7) increase |  |  |
| Ischemic stroke | 2588 | DOAC | 0.841 (0.794 – 0.891) | 15.9% (10.9-20.6) increase | 0.878 (0.820-0.940); p<0.001 | Supplemental materials A 6.6 |
|  |  | VKA | 1.027 (1.008 – 1.047) | 2.7% (0.8-4.7) decrease |  |  |
| All-cause death | 15,785 | DOAC | 0.982 (0.951 – 1.014) | NS | 0.956 (0.921-0.992); p=0.017 | Supplemental materials A 6.7 |
|  |  | VKA | 1.010 (0.982 – 1.038) | NS |  |  |
| Cardiovascular death | 6766 | DOAC | 0.984 (0.936 – 1.034) | NS | 0.974 (0.921-1.031); p=0.368 | Supplemental materials A 6.8 |
|  |  | VKA | 0.965 (0.944 – 0.986) | 3.5% (1.4-5.6) increase |  |  |
| Major bleeding | 6190 | DOAC | 0.931 (0.897 – 0.967) | 6.9% (3.3-10.3) increase | 0.965 (0.924-1.008); p=0.112 | Supplemental materials A 6.9 |
|  |  | VKA | 0.965 (0.944 - 0.986) | 3.6% (1.4-5.6) increase |  |  |

*HRs are relative hazard per 10% increase in PDC.

NS: not statistically significant; SSE: stroke or systemic embolism; TIA: transient ischemic attack; VKA: vitamin K antagonist; DOAC: direct-acting oral anticoagulant; PDC: proportion of days covered
